# Single-cell multiomic profiling of lung immune cells identifies novel asthma risk genes and cell-type specific functions

**DOI:** 10.64898/2026.02.05.26345013

**Authors:** Jing Gu, Donna C. Decker, Xiaoyuan Zhong, Anne I. Sperling, Carole Ober, Marcelo A. Nóbrega, Xin He, Nathan Schoettler

## Abstract

Genome-wide studies (GWAS) on asthma have identified nearly 200 genomic loci. However, the underlying mechanisms remain mostly elusive. While functional profiling of blood immune cell types has helped interpret asthma GWAS signals, high-resolution functional genomic data of lung immune cells, which differ from circulating immune cells, are lacking. We thus profiled single-cell multi-omics (RNA-seq and ATAC-seq) on lymphocytes of lung and spleen tissues from 9 donors. Cross-tissue comparison identified distinct transcriptomes for each immune cell type, but subtle differences in chromatin accessibility. We next assessed open chromatin regions (OCRs) of lung vs. blood, using a public dataset, for their enrichment of asthma risk. Strikingly, lung T cells showed unique contributions to heritability of adult-onset (AOA) and childhood-onset asthma (COA), beyond blood T cells. Using lung OCRs and previously fine-mapped variants for AOA and COA, we identified 43 cis-regulatory elements (CREs) likely contributing to asthma risk. By creating enhancer-gene maps from our single-cell data, we identified target genes for these CREs. We highlighted *CCR4* and *LRRC32* with their CREs displaying cell-type specific regulatory activities. Lastly, we built cell-type level gene regulatory networks (GRNs) to identify target genes of transcription factors (TFs). Lung GRNs not only shed light on the cell-type specific functions of several TFs that are known asthma risk genes, but also allowed us to detect novel TFs such as *STAT1* that may regulate asthma-related biological pathways in CD4 T cells. Our results demonstrate the utility of single-cell multiomics to identify asthma risk genes and understand their cell-type specific functions.

## 1 Introduction

Asthma is a common lung disease, typically characterized by chronic airway inflammation. Both innate and adaptive immune cells are implicated in asthma pathogenesis, including dendritic cells (DCs), eosinophils, neutrophils, lymphocytes, innate lymphoid cells (ILCs), and mast cells (Chou and Li, 2018; Hammad and Lambrecht, 2021). These immune cell types interact with neighboring tissue cells such as epithelial and smooth muscle cells, which also contribute to asthma pathogenesis. The cause of asthma Iis therefore complex, with genetics playing an important role.

Asthma is a heritable disease. The estimated heritability of asthma ranges from 35% to 95% (Ober and Yao, 2011). With common variants, it was estimated that 10-33% of asthma risk, depending on the age of onset, is due to genetic variation (Pividori et al., 2019). Mapping genetic variants associated with asthma risk would allow us to identify important genes involved in asthma-related biological processes and point out potential therapeutic targets. Collectively, genome-wide association studies (GWAS) of asthma have identified *>*150 significant loci, but due to linkage disequilibrium (LD), the causal variants in these loci are often unknown (Pividori et al., 2019; Tsuo et al., 2022). As in most other common traits, the majority of asthma-associated variants are in non-coding regions. With no direct consequences on proteins, disease-associated variants have been found to often play regulatory roles in disease (Finucane et al., 2015; Maurano et al., 2012). A major challenge of asthma genetics is thus to identify asthma risk variants, characterize their regulatory effects, and understand how dysregulation of the risk genes may lead to asthma.

To study regulatory functions of variants and genes, a common strategy is to leverage the epigenome and transcriptome in disease-relevant cells. Several studies, including our previous work, have shown that open chromatin regions in lymphoid cells are enriched for asthma-associated variants (Calderon et al., 2019; Zhong et al., 2025). However, most existing studies used epigenomic data from blood immune cells. Multiple lines of evidence suggest that lung immune cells provide a more relevant biological context to study asthma genetics (Masopust and Soerens, 2019; Takamura et al., 2019; Turner et al., 2014). Memory lymphocytes play important roles in the persistent conditions of asthma for their properties of being antigen-specific and long-lasting. In humans, memory lymphocytes from the lungs consist mostly of tissue-resident cells that do not circulate in the blood or secondary lymphoid organs. Previous studies have found abundant tissue-resident immune cells in lungs with distinct transcriptomic features (Purwar et al., 2011; Schoettler et al., 2019). Immune cells that reside in other tissues like adipose and skin have shown importance in tissue homeostasis, function, and immune surveillance (Heath and Carbone, 2013; Lu et al., 2019). Given their unique properties and important functions, we thus hypothesized that lung-resident immune cells play more direct roles in asthma genetics. To our knowledge, no studies have systematically examined the contribution of lung-resident immune cells to the genetic risk of asthma, beyond those from blood immune cells.

In this study, we profiled single-cell multiomics (gene expression and chromatin accessibility) on lymphocytes from the lung (Fig. 1a). For comparison, we also profile paired spleen samples from the same donors. This dataset allowed us to map open chromatin regions (OCRs) in multiple subsets of lung immune cells, and assess the contribution of regulatory sequences in each immune cell type to the genetics of asthma. By combining our data with published fine-mapping results of asthma, we identified novel asthma-associated cis-regulatory elements, nominated the target genes of asthma risk variants and their relevant cellular contexts.

**Figure 1:**
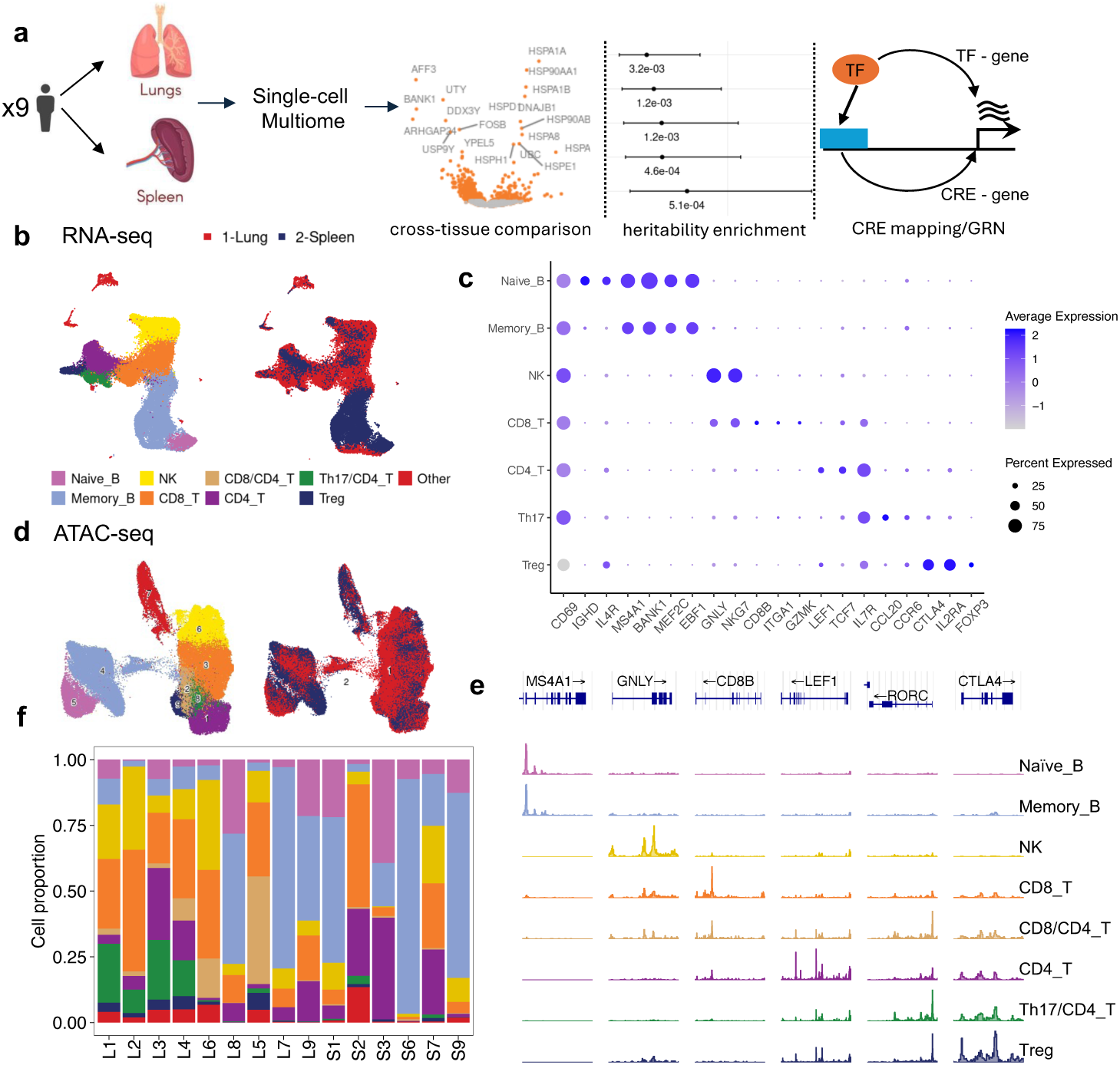
Characterization of immune cell populations in lung and spleen. **a**, An overview of experiment design and computational analyses. **b**, UMAP for the pooled RNA-seq libraries (N = 53647), colored by cell type (left) or tissue of origin (right). **c**, Dot plot for expression levels of *CD69* and other known marker genes for immune cell types. **d**, UMAP for the pooled ATAC-seq libraries (N = 102277), colored by cell type or tissue of origin. **e**, Track plots for chromatin accessibility within 10Kb of TSS of known marker genes - *MS4A1* (B cells), *GNLY* (NK cells), *CD8B* (CD8 T cells), *LEF1* (CD4 T cells), *RORC* (Th17 cells), *CTLA4* (Treg cells). **f**, A barplot of cell type compositions by donor and tissue sample. Colors represent different cell types as shown in fig. 1d. Donors were labeled by L (lung) or S (spleen) and the labels like L1 and S1 come from the same donor.

Lastly, we took advantage of our unique dataset to address another important gap in our understanding of asthma, namely, how dysregulation of gene expression through transcription factors (TFs) may lead to asthma. TFs are key regulators of immune cell states (Hosokawa and Rothenberg, 2021; Monticelli and Natoli, 2017). Several known asthma risk genes are TFs. What are the genes these TFs regulate and what cell types these TFs exert their effects on in asthma are largely unknown. Using our single-cell multi-omic data, we built gene regulatory networks (GRNs) linking TFs with their target genes. We used these GRNs to understand the functions of asthma TFs and identify additional TFs that may control important asthma-related biological processes.

## 2 Results

### 2.1 Single-cell profiling of gene expression and chromatin accessibility in lung and spleen immune cells

Single-nucleus multi-omic profiling (snRNA-seq and snATAC-seq) was performed in lung and spleen leukocytes from organ donors whose organs were not used for transplantation (n = 9). After read mapping, filtering, and data integration, we identified 53,647 cells and 8 clusters by gene expression (Supplementary Fig. 1). We used the gene expression data to annotate the main types of lymphocytes - naive B cells (n = 2884), memory B cells (n = 15794), NK cells (n = 8531), CD4 T cells (n = 7866), CD8 T cells (n = 12631), as well as a small number of “Other” cells (Fig. 1b). We further annotated subpopulations of CD4 T helper cells as regulatory T (Treg) cells and T helper 17 (Th17) cells (Fig. 1b). The patterns of known marker genes for these cell types confirmed these annotations: *MS4A1*, *BANK1* are highly expressed in B cells, *GNLY*, *NKG7* in NK cells, *CD3D*, *IL7R* in T cells, *CCL20* in Th17 cells and *FOXP3*, *CTLA4* in Treg cells (Fig. 1 c). We also evaluated the expression of CD69, a marker of tissue-resident lymphocytes. Approximately 75% of cells express high *CD69* across almost all the cell types (Fig. 1c), confirming that the majority of cells are tissue-resident. Our snATAC-seq libraries, after filtering, had more cells than RNA-seq libraries (Supplementary Fig. 2). In total, we had 102,277 cells with ATAC libraries. We used ArchR (Granja et al., 2020) for dimensional reduction and iterative clustering. To annotate the cell types from ATAC-seq libraries, we identified 9 major clusters and transferred annotations from matched RNA-seq data, using a majority voting approach for each cluster (Method). For two clusters, there is some ambiguity in assigning the dominant cell type, thus we annotated them with dual cell types: Th17*/*CD4 T and CD8/CD4 T (Supplementary Fig. 3). Using the uniform manifold approximation and projection (UMAP) of ATAC-seq data, we observed that cells cluster together according to the main types of lymphocytes (Fig. 1d). We called peaks in each cell type separately for lung and spleen with MACS2 (Yong Zhang et al., 2008). We found 70-122K peaks (501 bp) per cell type, with the union of ∼250K peaks. We further confirmed cell types by comparing the chromatin landscape at known marker genes. The combined track plots clearly demonstrate higher chromatin activity for marker genes of the corresponding cell type (Fig. 1e). For example, peaks in the gene region of *MS4A1* were observed in naïve B cells and memory B cells, but not in other cell types. Lastly, we separated cells by both donor and tissue and then examined cell type proportions in each group, which display extensive heterogeneity (Fig. 1f). Altogether, we generated a high quality single-cell based atlas of chromatin accessibility and gene expression in lung immune cells.

### 2.2 Lung-specific transcriptomic and epigenetic features

We assessed the difference of the epigenome and transcriptome of lung immune cells compared with spleen immune cells. We first observed differences in cell compositions by tissue site: T cells were the most abundant cell type in the lung and B cells were the most abundant in the spleen (Fig. 1b, f). We then performed differential expression (DE) analysis between lung and spleen, one cell type at a time. Memory B cells had the most DE genes (1027) and Naive B cells had the fewest (80) (Fig. 2**a**, Supplementary Table 2). To characterize these cross-tissue DE genes, we performed gene set enrichment analysis. Overall, DE genes showed broad enrichment of immune-related pathways across all cell types. Particularly, genes with higher expression in lung Memory B cells showed 8-10 fold enrichment for interleukin-2 and interleukin-4 production, which is essential for driving lung inflammation (Fig. 2b, Supplementary Table 3). These up-regulated genes in lung memory B cells were also strongly enriched for genes involved in T cell activation. Similar analysis was performed for the up-regulated genes in the spleen, but we found few enriched GO terms - immune response signaling pathway and B cell activation (Supplementary Fig 5, Supplementary Table 4). This suggests that gene expression regulation in lung memory B cells and T cells is distinct from those cell types in the spleen. Besides immune-related pathways, up-regulated genes across lung immune cell types are consistently enriched for genes involved in the processes related to protein folding including response to temperature stimulus. This shared feature reflects a conserved adaptive response to the lung microenvironment (Fig. 2b, Supplementary Table 3).

**Figure 2:**
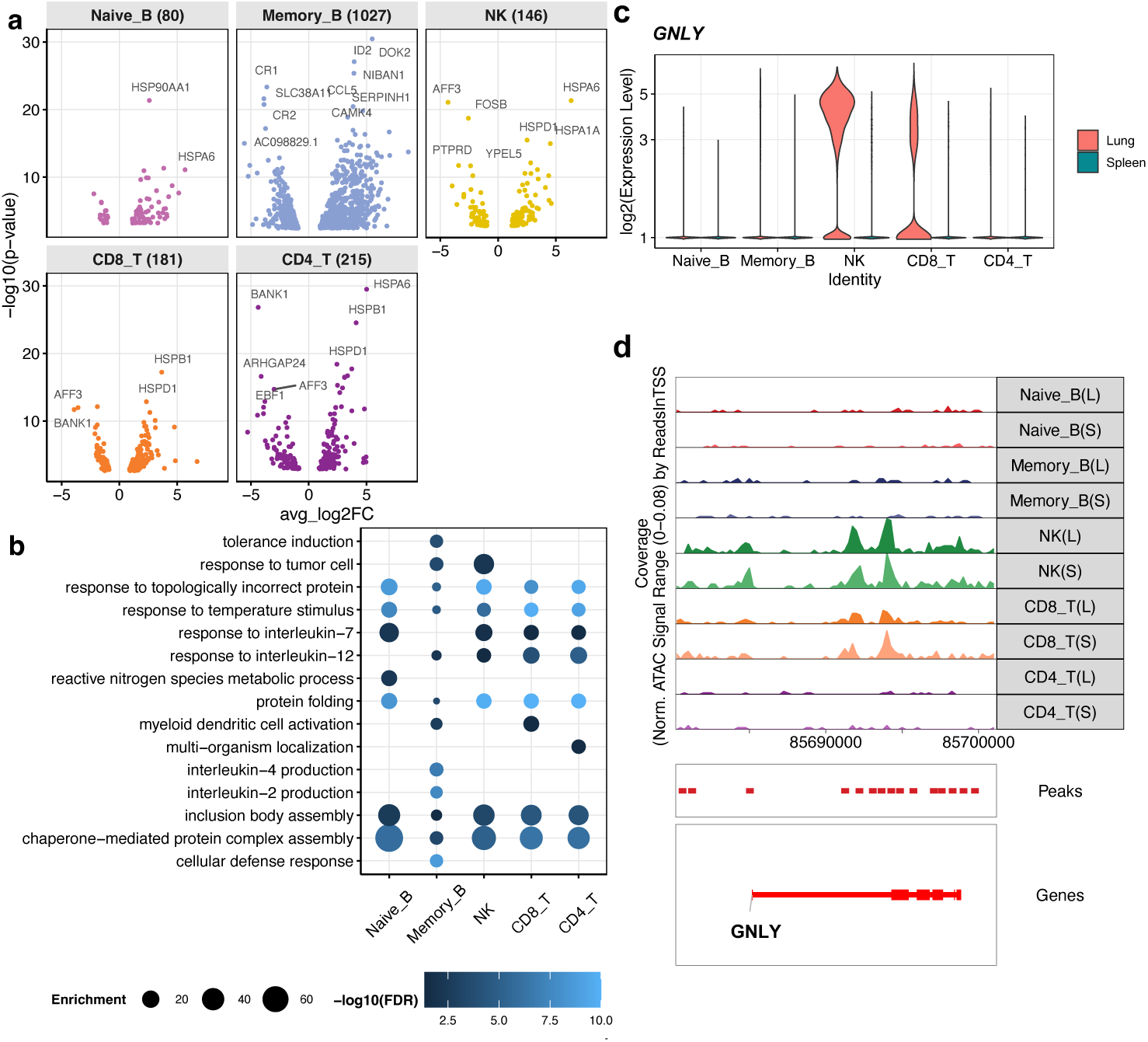
Cross-tissue comparison at both transcriptomic and epigenetic levels by cell types. **a**, Volcano plots for differential gene expression in log_2_ fold change (x-axis) across 5 immune cell types. Genes with higher expression in the lung have positive values. The numbers of differentially expressed (DE) genes were shown in the parenthesis in the labels of each strip. **b**, Top Gene Ontology terms enriched for lung up-regulated genes across cell types. Bubble size denotes fold of enrichment, while color denotes -log_10_(FDR). **c**, Violin plots of GNLY expression across cell types separately for lung and spleen. **d**, Normalized chromatin accessibility tracks for *GNLY* locus across cell types in lung (L) and spleen (S).

Having established robust differences in gene expression between lung and spleen immune cell types, we then tested for differences in chromatin accessibility between these tissues. To confirm that we can identify differential accessibility, we first compared chromatin accessibility across cell types within each tissue. As expected, we were able to detect tens of thousands of differentially accessible peaks between cell types at a comparable sample size (Supplementary Table 5). In contrast, the cross-tissue comparison, detected relatively few differentially accessible peaks across all cell types (0 in Th17/CD4 T to 73 in Treg) (Supplementary Table 5). This suggests that chromatin accessibility pattern across tissues may be subtle, and that the differences in gene expression between lung and spleen is not primarily mediated by changes in chromatin accessibility. As an example, *GNLY*, encoding for granulysin protein, produced mostly by cytotoxic immune cells, has significantly higher expression in NK cells in the lung (Fig. 2c, Supplementary Table 5). However, the epigenetic tracks for *GNLY* showed overall similar accessibility between lung and spleen across cell types (Fig 2d).

While the differences in chromatin accessibility seemed small between lung and spleen immune cells, the transcriptome comparison supported the differences of lung immune cells from spleen. Thus using genomic data from lung immune cells may provide a more relevant dataset for interpreting genetic findings of asthma.

### 2.3 Chromatin accessibility landscape of lung immune cells sheds light on the genetic architecture of asthma and allergic diseases

Using chromatin accessibility data of lung immune cells, we assessed the contribution of each cell type to the genetics of asthma. Given that previous studies of asthma often used circulating immune cells to identify likely regulatory variants, we first compared the open chromatin regions (OCRs) from lung in our study with those from blood immune cells. For the blood data, we used OCRs from a published scATAC-seq dataset (Benaglio et al., 2023). The number of OCRs in lung immune cells ranged from 70K to 120K, with 25-35% not shared with blood (Fig. 3a). Then we applied stratified linkage disequilibrium score regression (S-LDSC) to assess the enrichment of heritability (*h*^2^) of asthma in OCRs by testing each cell type separately (Finucane et al., 2015). We considered GWAS of two subtypes of asthma, adult-onset asthma (AOA) and childhood-onset asthma (COA) from the UK Biobank, as earlier studies showed they may involve different cell types and mechanisms (Pividori et al., 2019; Zhong et al., 2025). For comparison, we also included GWAS of allergy (a combined phenotype of asthma, hayfever, and eczema) (Ferreira et al., 2017), height, and body-mass-index (BMI) (Yengo et al., 2018). Consistent with previous findings, we observed individual immune cell types (T and NK) in lung and blood showing significant enrichment for asthma and allergy, but not height or BMI (Fig. 3b).

**Figure 3:**
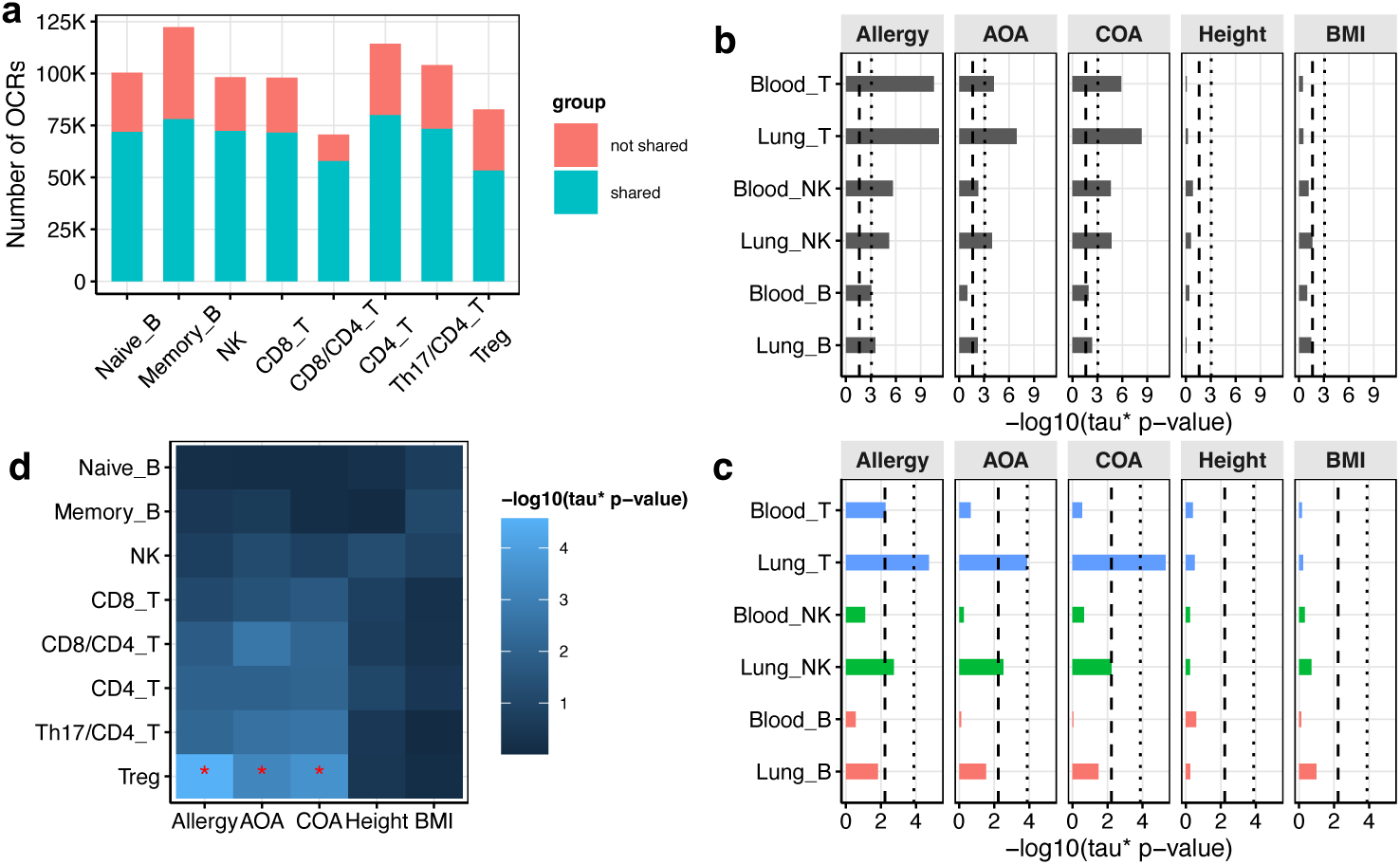
Enrichment of heritability of several traits in open chromatin regions (OCRs) from lung immune cells. **a**, Comparison of the numbers of OCRs from our data and those from a published blood scATAC-seq dataset. **b**, Barplot for -log_10_(Tau* p-values) of testing individual cell type of lung and blood with S-LDSC. The dash line corresponds to the FDR threshold at 0.05, while the dotted line corresponds to the Bonferroni threshold at 0.05. **c**, Barplot for -log_10_(Tau* p-values) of jointly testing paired tissues of individual cell type. The color scheme for the pairwise tests on T, NK and B cells were blue, green and red. **d**, Heatmap for -log_10_(Tau* p-values) of jointly testing all the lung immune cell types with a union set of OCRs of all the lung and blood cells. Tau* p-values below the Bonferroni cutoff at 0.05 were labeled with red asterisks.

Given that lung and blood OCRs show substantial sharing, we next aimed to tease out the unique contribution due to lung immune cells to asthma heritability. To do this, we jointly tested OCRs of paired lung and blood cells, one cell type at a time. Using the tau* coefficient from S-LDSC, this analysis effectively used the shared OCRs between lung and blood as “baseline”, and assessed the enrichment due to the tissue-specific component. We found that lung T cells showed much stronger enrichment in asthma and allergy than blood T cells. These results thus highlighted that lung immune cells made unique contributions to genetics of asthma, beyond the circulating immune cells.(Fig. 3c)

Lastly, we compared the relative contribution of different immune cell types for their genetic contributions. We performed joint analysis of all lung immune cells in S-LDSC, using the union of peaks from all blood and lung immune cells as background. We found that subsets of CD4 T helper cells, particularly Treg, were significantly enriched in heritability of asthma and allergy (Fig. 3d).

### 2.4 Single-cell transcriptome and epigenome informs the cell type-dependent functions of asthma risk variants

Recent work, including ours, has used statistical fine-mapping to identify putative causal variants of asthma (Clay et al., 2022; Saarentaus et al., 2025; Zhong et al., 2025). The regulatory functions of these variants, however, are often unknown. One approach is to assign variants to the nearest gene, but long-range gene regulation is common, and as a result, the true target gene(s) of a variant may not be in close proximity. Additionally, the relevant cell type where the asthma variants manifest regulatory effects are often unknown.

We started by obtaining a candidate list of cis-regulatory elements (CREs) that contain asthma risk variants. Our earlier fine-mapping study of asthma has prioritized 35 candidate CREs by intersecting fine-mapped variants with regulatory annotations in blood immune cells and several other asthma-related cell types including epithelia cells, fibroblast and smooth muscle cells (Zhong et al., 2025). Using our lung OCR map, we prioritized an additional 8 CREs that contain high confidence fine-mapped asthma variants, but missed in the earlier work (Methods). In total, we had 43 candidate asthma CREs.

Taking advantage of single-cell multiomics, we linked these candidate CREs to their target genes and assessed cell-type specific activity of the CREs. We used a recently developed method, scE2G (Sheth et al., 2024). This method predicts enhancer-to-gene links (E2G) using multiple features, such as activity by distance, Kendall correlations between chromatin and gene expression across single cells. We chose to use scE2G as it takes advantage of single-cell data, and can potentially discover cell-type specific target genes. Using scE2G, we identified target genes of 29 CREs, with 1-3 genes linked per CRE (Fig. 4**a**).

**Figure 4:**
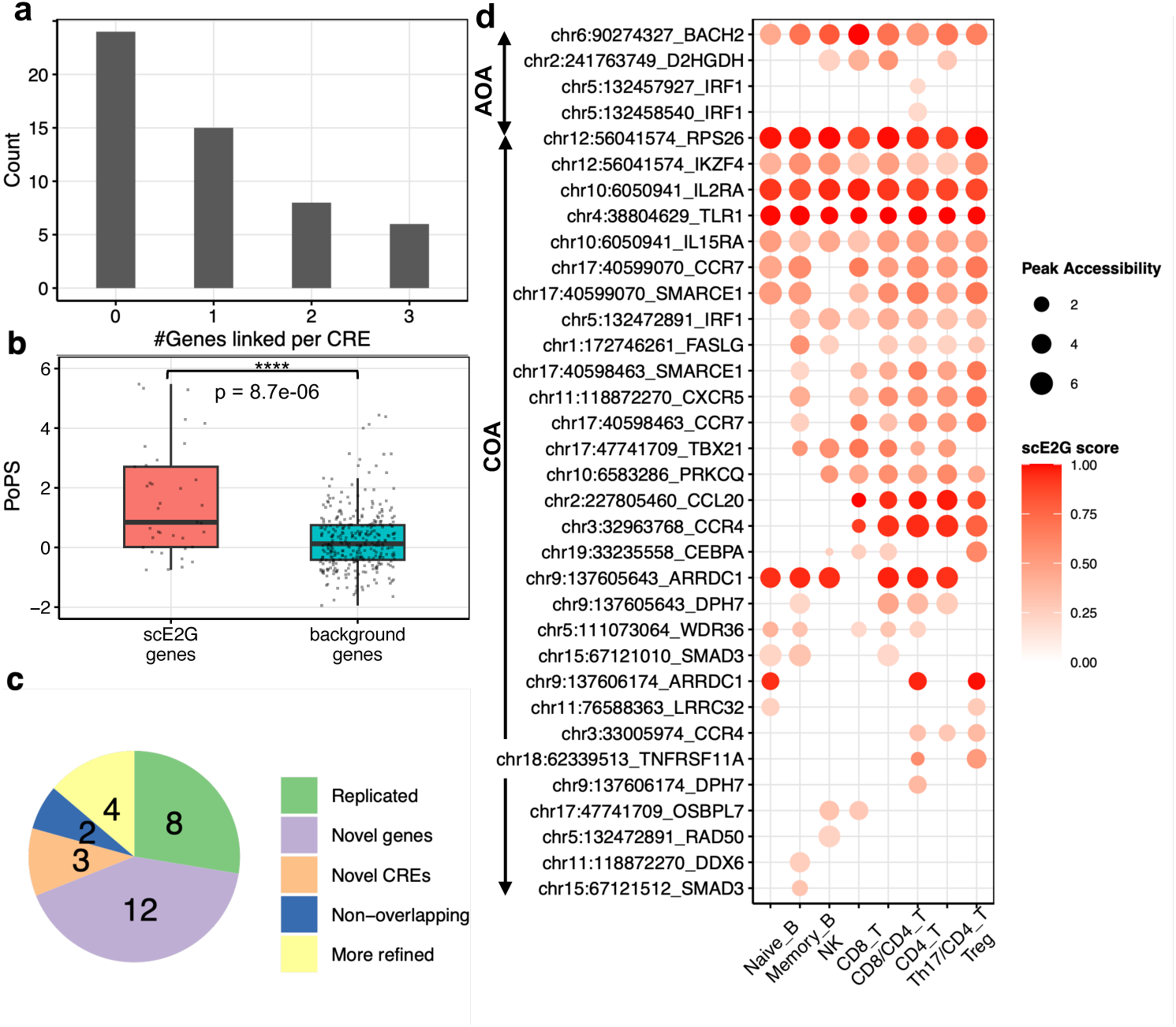
Asthma relevant cis-reglulatory elements (CREs) and their target genes in lung immune cells. **a**, Histogram showing the numbers of genes linked to CREs with scE2G multiome model. **b**, Comparison of polygenic priority scores (PoPS) for genes linked by CREs and those within 500KB of each CRE as control. The reported p-value was from Wilcoxon Rank-Sum Test. **c**, Pie chart for 29 linked CREs comparing our nominated risk genes with the prior publication. A locus is classified into five categories based on the nominated genes of the two studies. More refined: our study narrowed down candidate genes. Novel CREs: our study found new asthma CREs. Novel genes: our study found new genes from the same CREs. **d**, Bubble plot of the asthma CREs and the E2G supports in each cell type. Each row shows a CRE-gene pair. The size of the bubble denotes absolute peak accessibility and the color indicates scE2G prediction scores. CREs for adulthood on-set asthma are on the top and those for childhood on-set asthma are at the bottom. For simplicity, the CRE-gene pairs with genes in the top 25% expression quantile are shown (See the full list in Supplementary Fig. 6).

To validate these target genes as potential asthma risk genes, we leveraged polygenic priority score (PoPS). PoPS used GWAS data of a disease and many gene features such as tissue-specific expression patterns and pathway annotations to assess how likely a gene is a risk gene of the disease(Weeks et al., 2020). We compared PoPS of scE2G-nominated genes with the genes in the same region of CREs (Methods). Indeed, scE2G nominated genes show significantly higher PoPS than nearby genes based on the Wilcoxon ranksum test (p-value = 8.7e*^−^*^6^). (Fig. 4**b**, Supplementary Table 6). This result thus supported our identified target genes as likely risk genes for asthma.

Our earlier work also nominated target genes of asthma-associated CREs, using several criteria, such as distance, chromatin loop and eQTLs. The functional annotations used in this work were primarly from blood immune cells or other lung structural cells. We asked how our nominated target genes compared with these earlier results at each locus (Fig. 4c, Supplementary Table 7). In 8 loci, our nominated genes agreed with the previous findings (denoted as “replicated” in Fig. 4c). In 4 loci, we narrowed down to fewer linked genes (“more refined” in Fig. 4c). For example, one CRE was previously linked to both *SMAD3* and *MAP2K5*. Our approach nominated only *SMAD3*. *SMAD3* is a key transcription factor in the TGF-beta signaling pathway, which plays a central role in airway remodeling and immune regulation in asthma (Anthoni et al., 2007; Panek, Karbownik, Górski, et al., 2022). In contrast, *MAP2K5* has low PoPS score and we did not find strong literature evidence supporting this gene (Supplementary Table 6). Thus our approach likely identified the true target gene for this CRE.

In 12 loci, we discovered one or two additional linked genes (Fig. 4c, Supplementary Table 7). For example, scE2G predicted *IL15RA* to be linked to a CRE near *IL2RA*. This gene was plausible for its high PoPs score and its important role in lymphocyte homeostasis (Supplementary Table 6) (Rowley et al., 2009). Several other genes such as *ERBB3* and *RAD50* may indirectly modulate airway inflammation. Additionally, we were able to assign target genes to three out of eight novel asthma CREs that were missing in earlier work. The target genes include some novel candidate such as *WDR36*, which is critical for ribosome biogenesis and may potentially impact T cell activation (Skarie and Link, 2008). We evaluated all the genes nominated across all CREs through literature search. We found that the majority of them have asthma-related functions, but for many of these genes, direct links to asthma have not been established (Supplementary Table 6). Thus, our study nominated many potentially novel yet plausible candidates.

To evaluate the cellular contexts of asthma-associated variants, we next examined whether the predicted E2G links are shared across cell types or unique to a particular cell type. Using a heatmap to show the E2G links across cell types, we found they generally fall into several patterns: (1) broadly shared in all the cell types (eg. *BACH2* -linked CREs); (2) shared among all T cells (eg. *CCL20* -linked CREs); (3) shared among CD4 T cell subsets (eg. *TNFRSF11A*-linked CREs, Fig. 4d); (4) specific in certain cell type(s) (e.g. *SMAD3* -linked CRE in naive B cells). Overall, we observed more E2G links in CD4 T cells, consistent with the heritability enrichment results that we observed earlier in this study (Supplementary Fig. 6).

Altogether, our analyses highlighted novel CREs involved in asthma risk, refined the target genes of asthma risk variants, and shed light on cellular contexts where the candidate CREs act on the asthma risk. Below we highlighted two examples, with the E2G links found in CD4 T cell subsets.

### 2.5 Asthma candidate risk variants act through CD4 T cell-specific regulatory elements

Our first example is an COA-associated locus on chromosome 3. The region has several SNPs in LD associated with COA risk. Functional fine-mapping analysis from our earlier study identified two credible sets, with each set containing a distinct causal variant (Zhong et al., 2025). The SNPs in the credible sets overlap with two separate CREs, one in the intergenic region downstream of *CCR4*, one in the intronic region of *GLB1* (Fig. 5a). Both CREs had much higher chromatin accessibility levels in CD4 T cell subsets than other cell types and both were predicted to regulate the expression of *CCR4* (Fig. 5a). Interestingly, for the CRE located in *GLB1*’s intron region, the scE2G analysis highlighted *CCR4* as its target gene. Indeed, *CCR4* expression is specific to lung CD4 T cell subsets, while *GLB1* expression is largely similar across all lymphocytes (Supplementary Fig. 7). Our analysis thus supported *CCR4*, instead of *GLB1*, as the target gene for this CRE. CCR4 is a chemokine receptor highly expressed on Th2 cells, which play central roles in the initiation and persistence of airway inflammation. Studies in allergic asthma patients have found that the majority of airway T cells express CCR4 upon allergen challenge (Yang Zhang et al., 2017). By targeting CCR4 in mouse models, several studies have had success in attenuating allergic lung inflammation (Honjo et al., 2013). The convergence of two CREs to the same target gene boosts our confidence in *CCR4* as a potential mediator to drive asthma risks through CD4 T cells.

**Figure 5:**
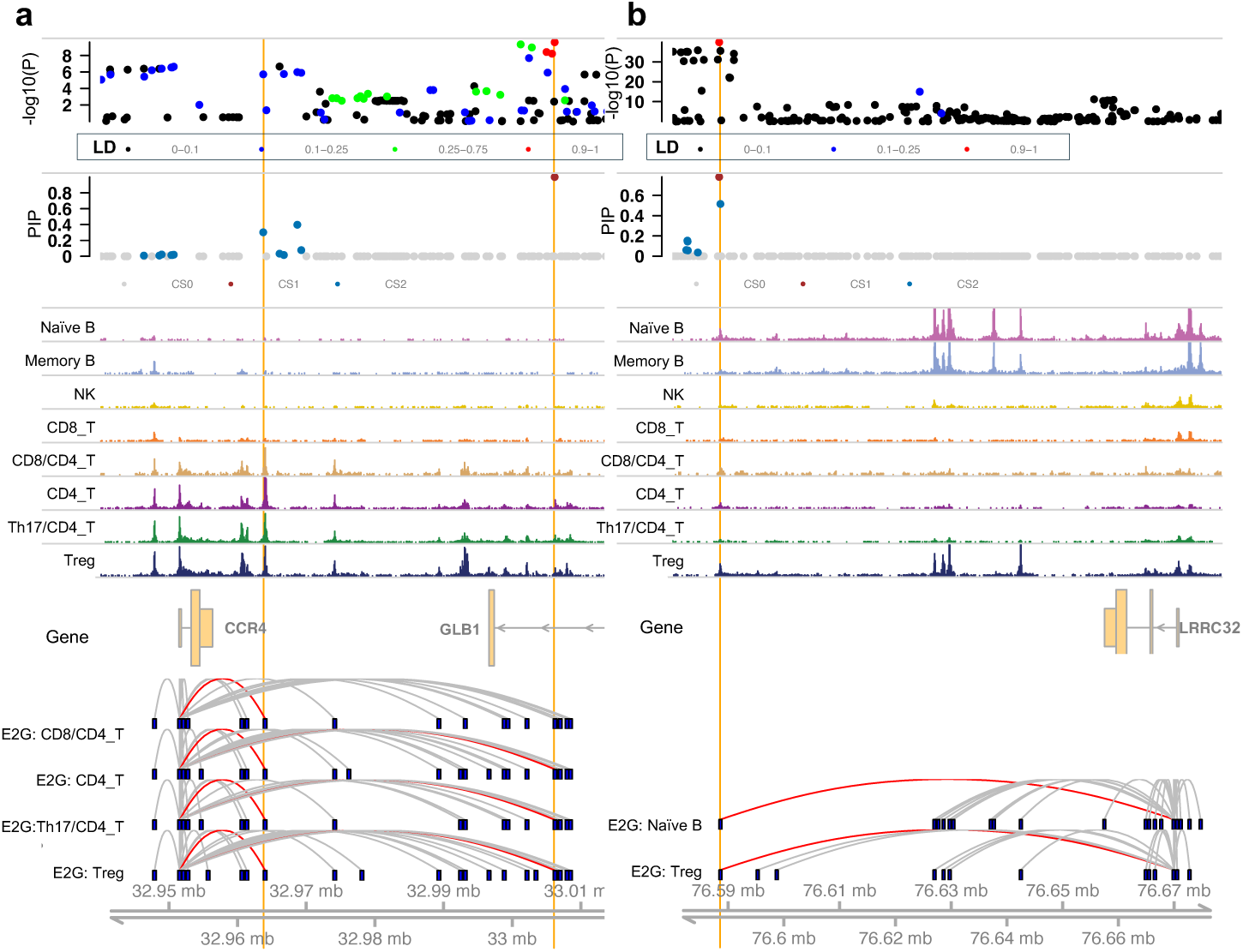
Two childhood-onset asthma related CREs show cell-type specific regulatory activity. From top to bottom, the first track shows the -log_10_ p-values from genetic associations. Each point is a SNP, colored by LD correlation with the top SNP ranked by p-value significance. The second track shows PIPs from fine-mapping. Each point is also a SNP, colored by credible set it belongs. CS0 means variants are not included in credible sets and thus no statistical significance to support them being causal, while credible sets CS1 and CS2 have at least 95% probability of containing one causal variant. The next eight tracks display chromatin accessibility from snATAC-seq of different lung immune cells. Below the gene track are scE2G predicted loops across lung immune cell types. The vertical bars represent candidate enhancers containing the fine-mapped SNPs and promoters of target genes. The loops from the candidate enhancers to the promoters of target genes are highlighted in red. **a**, At *CCR4* locus, two SNPs in two separate candidate enhancers are highlighted in orange, rs6795737 (left) and rs35570272 (right). **b**, At *LRRC32* locus, two high-confident SNPs, rs55646091 and rs11236797, overlap with the same CRE highlighted in orange.

The second asthma locus we examined is on chromosome 11 containing the gene *LRRC32* (Fig. 5b). Fine-mapping also identified two credible sets. The top SNPs from both credible sets, rs55646091 and rs11236797, respectively, overlap with the same CRE in the intergenic region downstream of *LRRC32*. This CRE has open chromatin in Naive B and regulatory T (Treg) cells, but is largely inaccessible in other lymphocytes (Fig. 5b). E2G analysis further linked this CRE to *LRRC32* in Treg as well as Naive B cells (Fig. 5b). Consistent with these observations, the expression of *LRRC32* is particularly high in Treg cells, and to a less extent in Naive B cells (Supplementary Fig. 7). Based on its roles in the development and function of Treg cells (Discussion), *LRRC32* is a plausible risk gene for asthma.

Our analysis of these two examples thus highlighted our ability to leverage single-cell multiome data from lung to annotate likely target genes of asthma risk variants and narrow down cellular contexts where the variants exert their effects.

### 2.6 Gene regulatory network analysis sheds light on the mechanisms of asthma-related transcription factors

By building gene regulatory networks (GRNs) in lung immune cells, we aimed to better understand the regulatory processes controlling cell-type specific expression patterns and gain insights into how these processes may be related to asthma. For our purpose, a GRN refers to the linking of transcription factors (TFs) and their target genes in each cell type. With the multiome dataset from lung, we built GRNs, one cell type at a time, using Dictys (Wang et al., 2023). This method predicts TF binding based on motifs and footprints from scATAC-seq data. The TFs bound to sequences near a gene would be considered candidate TFs for that gene. Dictys then associates the expression of each gene with the expression of candidate TFs using single-cell gene expression data, linking the TFs with their target genes.

We identified 252 TFs targeting 13,007 genes in our networks across 5 cell types. The 25th and 75th percentile for target gene counts per TF range from 165 to 212 (Supplementary Fig. 8a). Using the number of target genes of a TF in a cell type to define its regulatory activity in that cell type, we nominated TFs whose regulatory activities are cell-type specific (Fig. 6**a** “Marker TFs”). These include known lineage-specific TFs for B cells (e.g. *PAX5, SPIB, SPI1*) and T cells (e.g. *LEF1, FOXP3*). We also discovered novel lineage-specific TFs such as *ETV7* for Treg and *CEBPA* for CD8 T (Fig. 6**a**, see Discussion).

**Figure 6:**
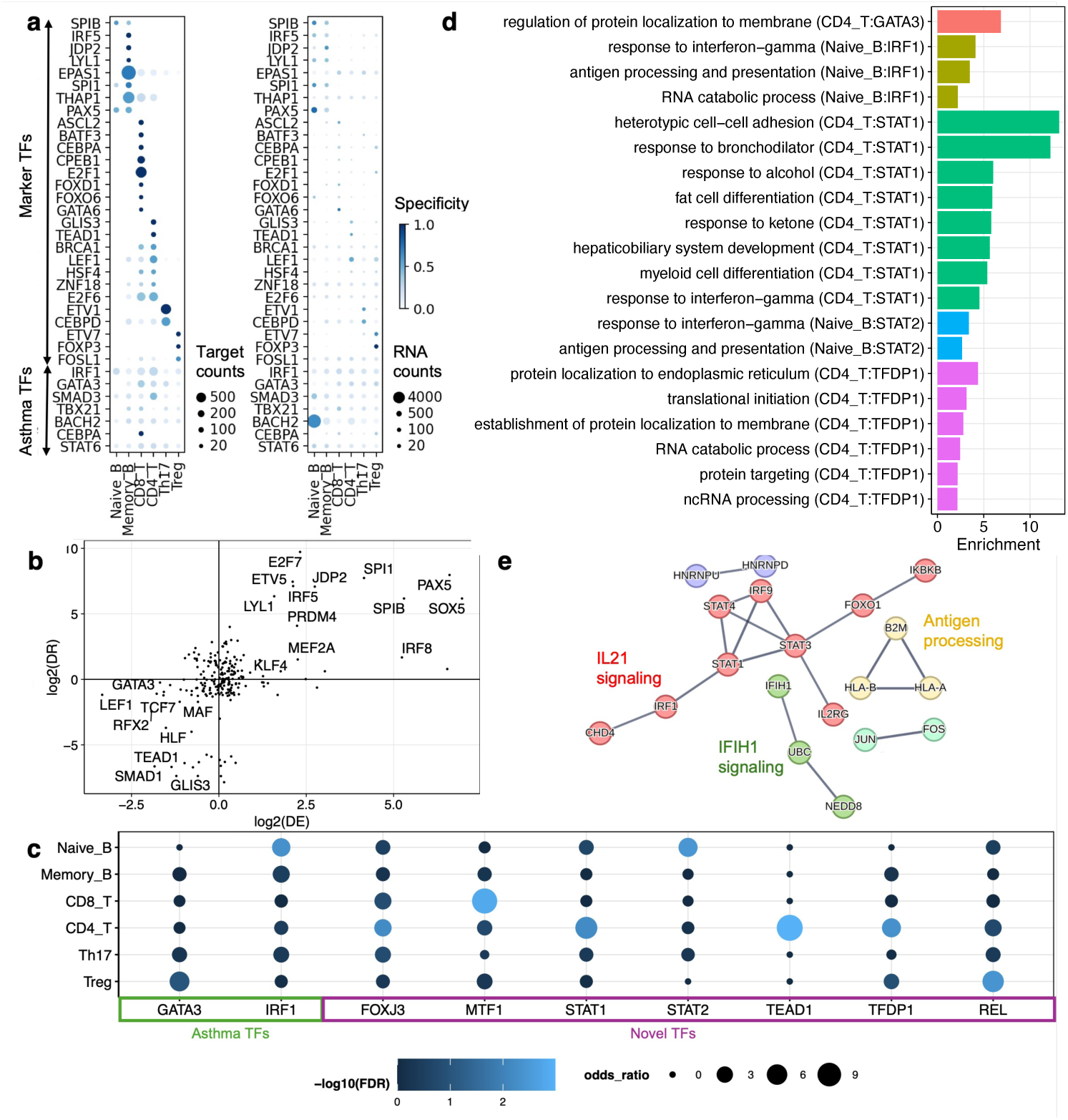
Cell-type level GRN analysis in lung immune cells. **a**, Heatmap for cell type specific TFs (“Marker TFs”) and asthma TFs (bottom), showing regulatory activity (left) and gene expression (right). Size of the data points denote target gene counts (left) or RNA read counts (right). Color represents specificity of the TFs across cell types (see Methods). **b**, Differential regulation (DR, Y-axis) and differential expression (DE, X-axis), at the log2 fold change scale, of TFs between B and T cell subtypes (Method). We chose the subtypes with comparable cell counts. Top TFs with at least 2 fold differences were labeled. A positive value of a TF means the TF targets larger number of genes and meanwhile displays higher expression in the subtypes of B cells than in those of T cells. **c**, Enrichment of asthma risk genes among target genes of candidate TFs in each lung immune cell type. Candidate TFs were grouped by previously nominated asthma risk genes (”asthma TFs”) in green and putative master regulators for asthma identified using our GRNs (”novel TFs”) in magenta. **d**, Enriched top GO biological processes terms for target genes of each candidate TF for asthma in the respective cell types that show significant enrichment in Fig 6c. (See the full list in Supplementary Fig. 11). **e**, Visualization of *STAT1* target genes in CD4 T cells from STRING network analysis. Genes with more than 0.9 confidence scores were shown, and several clusters were identified through k-means clustering: (1) interleukin-21 signaling (red), (2) Antigen processing and presentation (yellow), (3) Negative regulators of IFIH1 signaling (green), (4) Transcription factor AP-1 complex (light blue), (5) CRD-mediated mRNA stabilization (purple)

To better understand the TFs driving cell type differences, we compared two cell types, each from T and B cell subsets, by the difference of TF gene expression between the two cell types (differential expression), and the difference of the number of target genes (differential regulation). Known lineage-specific TFs including *PAX5* and *SPIB* tended to display large differences in both expression and target gene counts (Fig. 6b). We hypothesized that TFs with large differences using both metrics would be more likely to regulate genes that are differentially expressed between cell types. Indeed, 8 regulatory TFs specific to CD4 T cells (*>*=1.5 fold change using both metrics mentioned earlier), including *GATA3*, *LEF1*, *TCF7*, show significant enrichment for genes with higher expression in CD4 T cells than the B cell subset (Supplementary Fig. 8b). These results thus support the TFs we discovered as drivers of cell type differences in the transcriptome, and provided some validation of the GRNs in linking TFs to target genes.

We next used these GRNs to understand asthma genetics. Our previously nominated asthma risk genes include several TFs, yet we do not know how these “asthma TFs” affect other genes or pathways in the development of asthma. With GRNs of lung immune cells, we can now identify downstream genes for these TFs. Seven asthma TFs were found to have at least one target gene in our lung GRNs. Except *CEBPA*, these TFs display similar regulatory activities, i.e. comparable numbers of target genes, across cell types (Fig. 6**a**, “Asthma TFs”). Interestingly, we noticed that, even for the same TF, the actual regulatory networks across immune cell types are not identical and they are enriched in distinct pathways (Supplementary Fig. 9-10). For example, *STAT6* sub-networks across cell types show distinct top regulated genes, ranked by regulation effect sizes (Supplementary Fig. 9a). STAT6 target genes in regulatory T cells are uniquely enriched in pathways related to leukocyte proliferation, activation and cell-cell adhesion compared to other cell types (Supplementary Fig. 9b). These results thus highlighted the pleiotropic function of the asthma TFs, i.e. the same TFs may rewire to different genes under different cellular contexts.

Given the potentially different functions of the asthma TFs in different cell types, we hypothesize that their asthma-related function is likely mediated through biological processes in specific cell type(s). To test this, we asked whether the target genes of an asthma TF are enriched with known asthma risk genes in each cell type. Out of seven asthma TFs, *GATA3* targets in Treg cells and *IRF1* targets in Naive-B cells showed significant enrichment of asthma risk genes, but not in other cell types.(Fig. 6c, “Asthma TFs”). Gene set enrichment analysis reveals several biological processes enriched in the target genes of GATA3 and IRF1 in the respective cell types (Fig. 6d). These include important immune processes such as “response to INF-gamma” for IRF1 targets in B cells.

Having found the utility of GRNs in characterizing asthma TF targets, we searched for novel TFs that impact asthma-related genes or pathways. These TFs represent potential “master regulators” and can be potential targets for therapeutic interventions. To find such TFs, we tested the enrichment of asthma risk genes in each TF’s sub-network, one cell type at a time. In total, we nominated seven TFs with significant enrichment (FDR ≤ 0.15) in at least one cell type (Fig. 6c). For most TFs, their target genes showed enrichment of asthma risk genes only in single cell types. The target genes of these TFs were enriched in asthma-related biological processes (Fig. 6d). Notably, STAT1 targets in lung CD4 T cells include genes involved in multiple aspects of asthma, including T cell activation, myeloid cell differentiation, and response to bronchodilators, a common treatment for asthma (Fig. 6**d**, Supplementary Fig. 11). To further explore the functions of STAT1 target genes, we visualized *STAT1* sub-network of CD4 T cells with STRING (Szklarczyk et al., 2023). The sub-network contains several clusters of interacting genes. The main cluster involved *STAT1* and other genes that show enrichment in interleukin-21 signaling pathway, which promotes Th2 and other T cell inflammatory responses (Fröhlich et al., 2007) (Fig. 6e). The other clusters in the sub-network captured other related immune processes, such as regulators of early response genes (JUN-FOS), antigen presentation and processing (B2M and HLA genes) and viral sensing (IFIH1).

In summary, our results highlighted pleiotropic functions of TFs implicated in asthma, with potentially different functions in different lymphocytes, and identified novel candidate TFs regulating asthma-related processes. For most of these TFs, their asthma-related functions are likely limited to specific cell types. Examining target genes of these TFs in the relevant cellular context provided insights into their roles in asthma.

## 3 Discussion

In this study, we profiled single-cell epigenome and transcriptome of lung immune cells. We found that lung immune cells show distinct transcriptomes compared to spleen immune cells. OCRs across immune cell types show enrichment of asthma risk, with CD4 T subsets showing particularly strong enrichment. By integrating the E2G links inferred from our data with asthma fine-mapping, we nominated a number of putative risk genes as targets of the fine-mapped variants, highlighting two promising genes *CCR4* and *LRRC32*. Lastly, we built GRNs across the immune cell types. These networks shed light on the function of several asthma TFs and allowed us to nominate novel regulators of asthma-related biological pathways.

Our asthma CRE analysis highlighted *LRRC32* as a COA risk gene, with its asthma function likely mediated by Treg cells. *LRRC32* encodes the Glycoprotein A repetitions Predominant (GAPR), a membrane receptor for latent TGF-*β*. GARP may serve as a protective role by controlling the activation and surface presentation of TGF-*β*, which is essential for Treg cells to suppress inflammation and maintain immune tolerance (Zimmer et al., 2022). However, no functional studies have shown the direct effect of GARP on Treg-mediated airway responses in asthma. Our results, for the first time, have demonstrated *LRRC32* as the potential mediator for asthma genetic risk in Treg cells. These findings enable new hypotheses and opportunities around modulating the regulatory activity *LRRC32* in Treg cells.

Using GRN analysis, we highlighted two novel TFs as cell type specific regulators - *ETV7* in Treg and *CEBPA* in CD8 T cells (Fig. 6a). *ETV7* is a member of the ETS family of TFs, and several of which are known to play roles in immune cell differentiation (Yang et al., 2024). *CEBPA* is known to be a master regulator of the myeloid lineage, in particular, regulating granulocytic-monocytic differentiation (Pundhir et al., 2018; Radomska et al., 1998; P. Zhang et al., 2004). Supporting our finding of the role of CEBPA in CD8 T cells, a recent study found that up-regulating CEBPA with a small activating RNA activated CD8 T lymphocytes (Zhou et al., 2019). Thus, these two TFs are plausible candidates for regulating cell-type specific gene expression in lung immune cells. Using GRNs, we also investigated the networks of *STAT6* and *STAT1*, as potential master regulators for asthma. Both have been implicated in asthma pathogenesis: STAT6 promotes Th2-driven allergic inflammation characteristic of asthma and STAT1 participates in inflammatory processes in asthma with roles in antiviral immunity (Wang et al., 2023).

We note several limitations in our study. First, we acknowledge sample size is a limiting factor for our comparative analyses. This may lead to under-estimation of the lung-spleen difference. However, we are less concerned about comparing OCRs between lung and blood for T and NK cells because their number of cells and library quality were comparable. Second, the cell labels from about half of our ATAC-seq data were inferred due to lack of matched RNA-seq data. The majority-voting approach we took to infer cell types depends on the clustering of ATAC-seq data, and thus the uncertainty in assigning cell type for individual cells was not considered. Nonetheless, we were able to get an overall similar annotation for clusters compared to an alternative method (Supplementary Fig. 4). Thirdly, scE2G model uses ABC scores computed from distance rather than Hi-C data of relevant cell types as features. Cell-type specific chromatin loop data may improve scE2G’s accuracy on predicting target genes. Fourthly, our GRN analysis may be conservative, missing target genes of TFs. We found about 200 targe genes per TF, compared to often thousands of TF binding sites from typical ChIP-seq experiments or differentially expressed genes from TF knockout. Some steps of the GRN reconstruction have room for improvement. For example, the initial step of network construction relies on TF footprint detection using a published method. Newer methods for TF footprint discovery may uncover more footprints (Hu et al., 2025).

In summary, our study provided valuable data on lung-resident immune cells, helped identify asthma risk genes from the associated loci, and reconstructed GRNs that shed light on the genetics of asthma. Our approach of leveraging single-cell multiomics data with GWAS to identify risk genes through E2G analysis and cell-type level GRNs can be applied to a wide range of complex traits and diseases.

## 4 Methods

### 4.1 Human tissue procurement

Human lung samples were obtained from organ donors whose lungs were not used for transplantation through the Gift of Hope Regional Organ Bank of Illinois. Donors with *>*10 pack-years of tobacco use were excluded from this study. Because samples from this study were from deceased donors, they do not qualify as “human subjects” (determined by the Institutional Review Board at the University of Chicago).

### 4.2 Leukocyte isolation

Cells were processed as previously described. Briefly, lung tissue was perfused with sterile fetal bovine serum and phosphate-buffered saline. The right lower lobe was minced and digested. Separately, spleen and lung tissue mononuclear cells were enriched using density gradient centrifugation and cryopreserved.

### 4.3 Single-cell processing

Cells were thawed and resuspended in complete RPMI media and were rested at 37*^◦^*C for two hours prior to being processed by the University of Chicago Genomics Facility using 10XGe-nomics Multiome reagents. Sequencing was performed using NovaSeq X (See supplementary Table 1 for library information)

### 4.4 Pre-processing of 10x Multiome Data

The raw FASTQ files for all the multiome libraries were converted from base call files by the sequencing core at the University of Chicago. We examined the quality of sequencing reads with FastQC (v.0.12.1)(Andrews, 2010) and found 7 RNA-seq libraries did not pass QC criteria, and thus excluded from downstream analyses. Then we followed the cellranger-arc workflow (v.2.0.2) (10x Genomics, 2018) provided by 10x Genomics for preprocessing. The reference package needed to run the workflow was downloaded from 10x Genomics, including the human genome assembly FASTA file in GRCh38 and a version 32 GTF file of gene annotation. We further combined data from multiple libraries separately for lung and spleen tissue using cellranger-arc “aggr” function, with read depth normalized, and obtained an aggregated matrix for each tissue. For quality control at cell level, we followed Seurat workflow (v4.4.0) (Hao et al., 2021) to set up seurat objects for each tissue, and selected cells for further analysis with the following criteria - 1) Lung aggregate libraries: ATAC counts between 500 and 80K, RNA counts between 250 and 25K, mitochondrial reads less than 10%. 2) Spleen aggregate libraries: ATAC counts between 500 and 80K, RNA features between 250 and 4K, and mitochondrial reads less than 10%.

### 4.5 Integrating single-cell RNA-seq datasets

We performed two rounds of integrative analysis 1) across samples within each tissue and 2) across tissue, based on Seurat’s anchor-based RPCA approach. For each tissue, we implemented the following procedures to project the count matrix into low-dimensional space.

1. Normalizing the count matrix with Seurat::NormalizeData
2. Identifying variable genes with Seurat::FindVariableFeatures
3. Selecting features used for integration with Seurat::SelectIntegrationFeatures
4. Scaling the count matrix into zero mean and unit variance with Seurat::ScaleData
5. Running PCA with Seurat::RunPCA

Next, we selected top 20 dimensions and iteratively identified anchors that link cells across multiple scRNA-seq datasets within each tissue in the low dimension space with Seurat::FindIntegrationAnchors. With the identified anchors, we generated an integrated matrix for multiple samples of each tissue. This process was repeated for integrating datasets across tissues, which generates a final seurat object with 53647 cells and 187471 unique RNA molecules.

### 4.6 Dimension reduction, clustering and cell type annotation

We followed the standard workflow of Seurat on the integrated matrix of lung and spleen samples to identify cell clusters, which involves several steps as below.

1. Scaling the integrated matrix with Seurat::ScaleData
2. Running PCA to obtain top 30 principal components with Seurat::RunPCA
3. Identifying nearest neighbors at PCA space of 30 dimensions using Seurat ::FindNeighbors
4. Performing graph-based clustering with resolution level at 0.5 using Seurat::FindClusters
5. Visualizing clusters at low dimension with Seurat::RunUMAP

We further assigned cell types to each cluster with CellTypist (v1.7.1) (Dominguez Conde et al., 2022), an automatic cell type annotation tool for scRNA-seq datasets. We selected the model called “immune All Low.pkl”, for its comprehensive list of immune cells and enabled majority voting to assign the dominant cell type label for each cell type. To validate cell type annotation, we plotted expression of known marker genes for immune cells with Seurat::Dotplot.

### 4.7 Differential gene expression analysis between tissue

We performed differential gene expression between tissue with pseudo-bulk samples using DE-Seq2 (v1.38.3) (Love, Huber, and Anders, 2014). The test was done on the RNA count matrix aggregated by donor, tissue and cell types using Seurat::AggregateExpression. We also adjusted for covariates including batch, ethnicity, age, and sex.

### 4.8 Processing single-cell ATAC-seq

We processed all the single-cell ATAC-seq libraries with ArchR (v1.0.2) (Granja et al., 2020) workflow that involve the following steps:

1. Performing Initial QC at minTSS = 4 and minFrags = 1000 with ArchR::createArrowFiles
2. Detecting and removing doublets at k = 10, LSTIMethod =1, with ArchR::addDoubletScores and ArchR::filterDoublets
3. Filtering cells with low number of unique fragments from one sample named as COB-5 to top 3K so that QC metrics are more consistent across samples
4. Performing dimension reduction using iterative latent semantic index approach over genome-wide 500-bp tiles with ArchR::addIterativeLSI
5. Performing graph-based clustering at resolution = 0.8, using ArchR::addClusters
6. Visualizing data in low-dimensional space with ArchR::plotEmbedding

The final ArchR object with pooled samples from lung and spleen consists of 102277 cells, with the median unique number of fragments around 6K and the median enrichment of reads at transcription start site around 16.

### 4.9 Annotating cell clusters for single-cell ATAC-seq data

For single-cell multiomics datasets, we can directly annotate clusters for scATAC-seq data using labels from scRNA-seq. However, not all the cells have matched scRNA and scATAC sequencing data that pass quality checks for our data. We took two approaches to annotate cell type - majority voting and transfer labeling using ArchR::FindTransferAnchors. For majority voting, we assign the dominant cell type to each ATAC-seq cluster based on the cells in the same cluster that have labels from matched scRNA-seq. This approach should perform reasonably well, because for the majority of the clusters the labeled cells were dominated by one single cell type(Supplementary Fig. 3b). For two clusters (C14 and C16) with ambiguity in defining the dominant cell type, they were T cell subsets with overall similar chromatin accessibility. We simply retained them as their own cluster, named as the mixture of two cell types - CD8/CD4^+^T and Th17/CD4^+^T. The two approaches give overall similar annotations for each cluster (Supplementary Fig. 4), so we used the former approach to annotate cell types. With the satisfied cell type annotations, one library, named as SMO-10, was found to have mostly non-lymphocytes, so it was excluded from downstream analyses.

### 4.10 Creating pseudo-bulk replicates and Peak calling

We grouped cells by both tissue and cell type and followed the ArchR workflow to create pseudo-bulk replicates from each cell group with ArchR::addGroupCoverages. Peak calling was then performed for each defined cell group and was assessed for reproducibility with ArchR::addRreproduciblePeakSet. ArchR generated 501-bp-fixed-width peaks, which underwent iterative removal to retain the most significant one when multiple peaks are found to overlap with each other across cell groups.

### 4.11 Differential chromatin accessibility analyses

For identifying marker peaks for each cell type, we called ArchR::addMarkerFeatures on the peak matrix comparing one cell type against all others using wilcoxon rank sum test, adjusted by TSS enrichment and logarithm of fragment counts.

For identifying differentially accessible peaks between tissue, we first generated count matrix by donor, tissue and cell type and then tested peaks with differential accessibility using DESeq2. We also adjusted for covariates including batch, ethnicity, age, and sex.

### 4.12 Heritability enrichment test with Stratified-LDSC

To assess disease heritability explained by each immune cell type, we applied stratified LD-score regression (v1.0.1) (Finucane et al., 2015) on several GWAS datasets using functional annotations derived from open chromatin regions (OCRs) of lung and blood immune cells (Benaglio et al., 2023). To assess heritability enrichment, we tested individual cell type in lung with a union set of lung and blood peaks as well as 53 baseline annotations. For cross-tissue comparison, we jointly tested OCRs of paired lung and blood cells, one cell type at a time. We used the standardized effect size (Tau*) as defined by Kim et al. to quantify the effects that are unique to each focal cell type (Kim et al., 2024).

### 4.13 Identifying cis-regulatory elements (CREs) involved in asthma genetic risk

We identified 43 CREs containing fine-mapped asthma genetic variants. The CREs came from two sources. The first 35 CREs were from those prioritized in our earlier work with ePIPs ≥ 0.5 (Zhong et al., 2025). Here ePIP of a CRE means the sum of PIPs of all fine-mapped SNP(s) located in that CRE. These sequences were then overlapped with our OCRs by extended 250 bp. We further filtered out those *>*250 bp away from the fine-mapped variants (PIP≥0.3). This step ensures that the SNPs are contained in regulatory elements and potentially impact enhancer activity. The other 8 CREs came from directly overlapping our OCRs with fine-mapped variants (PIP≥0.3) by extended 250 bp.

### 4.14 Linking accessible peaks to genes

We linked putative enhancers to target genes using a machine-learning model from a recently developed tool called scE2G (v1.2) (Sheth et al., 2024). This tool allows us to leverage our partially matched sc-multiomics datasets to infer target gene(s) for each regulatory element in open chromatin identified per cell type. We ran the scE2G pipeline with the raw count matrix of scRNA-seq and fragment matrix of scATAC-seq per cell cluster as input and specified the predictive models - multiome data to assign scores to each enhancer-gene pair. To map peaks from scE2G pipeline to our OCRs, we required at least 250 bp overlapping between scE2G peaks and ours.

To further validate scE2G predicted genes, we computed polygenic priority scores (PoPS) for scE2G predicted genes and all genes within 250KB of CREs using GWAS of AOA due to its large sample size (Weeks et al., 2020). With the full set of gene features collected by PoPS (v0.2) (Weeks et al., 2020), we computed PoPS scores for each gene.

### 4.15 Reconstructing Gene Regulatory Networks (GRNs) of lung and spleen immune cells

We built gene regulatory networks (GRNs) using our partially matched sc-multiomics datasets with Dictys (v.1.1.0) (Wang et al., 2023). Separate GRNs were constructed for each subset of T and B cells from both tissues. We first inferred binary TF binding networks based on TF footprinting and motifs from single-cell chromatin accessibility. Then we refined the networks with single-cell transcriptomics data to associate gene expression with TF expression. The output of this step is the association strength between each gene and a set of TFs. This process results in cell-type level GRNs. To better interpret our GRNs, we set the network sparsity level to be 0.02 across all the cell types. When identifying cell-type specific TFs, denoted as “marker TFs”, we used the normalized target counts across TFs and cell types (i.e. specificity of the TFs) as defined in the Dictys paper. We also followed what the paper suggested to compare differential regulation (the differences in number of target genes) and differential expression (the differences in gene expression) between two cell types.

### 4.16 Data and code availability

Our lung single-cell multiomics data will be made available prior to publication of the manuscript. This study uses genotype and phenotype data from the UK Biobank under application number 44300. We obtained summary statistics of AOA and COA GWAS performed with UK Biobank version 3 genotypes from Zhong et al., 2025. The GWAS summary statistics for allergy were downloaded from https://genepi.qimr.edu.au/staff/manuelf/gwas_results/main.html. The GWAS summary statistics for height and BMI were downloaded from https://portals.broadinstitute.org/collaboration/giant/index.php/GIANT_consortium_data_files. The blood scATAC-seq data were downloade from https://zenodo.org/records/7375095.

The custom codes generated for this study will be available in https://github.com/xinhe-lab/Lung_scMultiomics_paper.

## Data Availability

All data produced in the present study are available upon publication

## 4.17 Acknowledgments

The authors would like to acknowledge Dr. Kevin Luo for assistance with the mapgen R package, Dr. Peter Carbonetto and Dr. Xiaotong Sun, for helpful discussions about computational analyses, and the AADCRC center at University of Chicago for all the feedback and support on the project. The authors gratefully acknowledge the Gift of Hope Organ and Tissue Donor Network and their donors and their families for donating tissues used for this study.

## 5 Author Contributions

J.G. processed the datasets, performed the analyses, interpreted the results, and prepared the manuscript; D.D. performed single-cell multiomics in lung and spleen samples; X.Z. performed fine-mapping analyses; D.D., A.I.S., C.O., M.A.N., X.H. and N.S. assisted with results interpretation; X.H. and N.S. supervised the computational analyses; X.H., A.I.S., and N.S. designed the study and interpreted the results. All authors contributed to writing the manuscript. All authors read and approved the final manuscript.

## Supplementary Information

### Supplemental Figures

**Figure S1:**
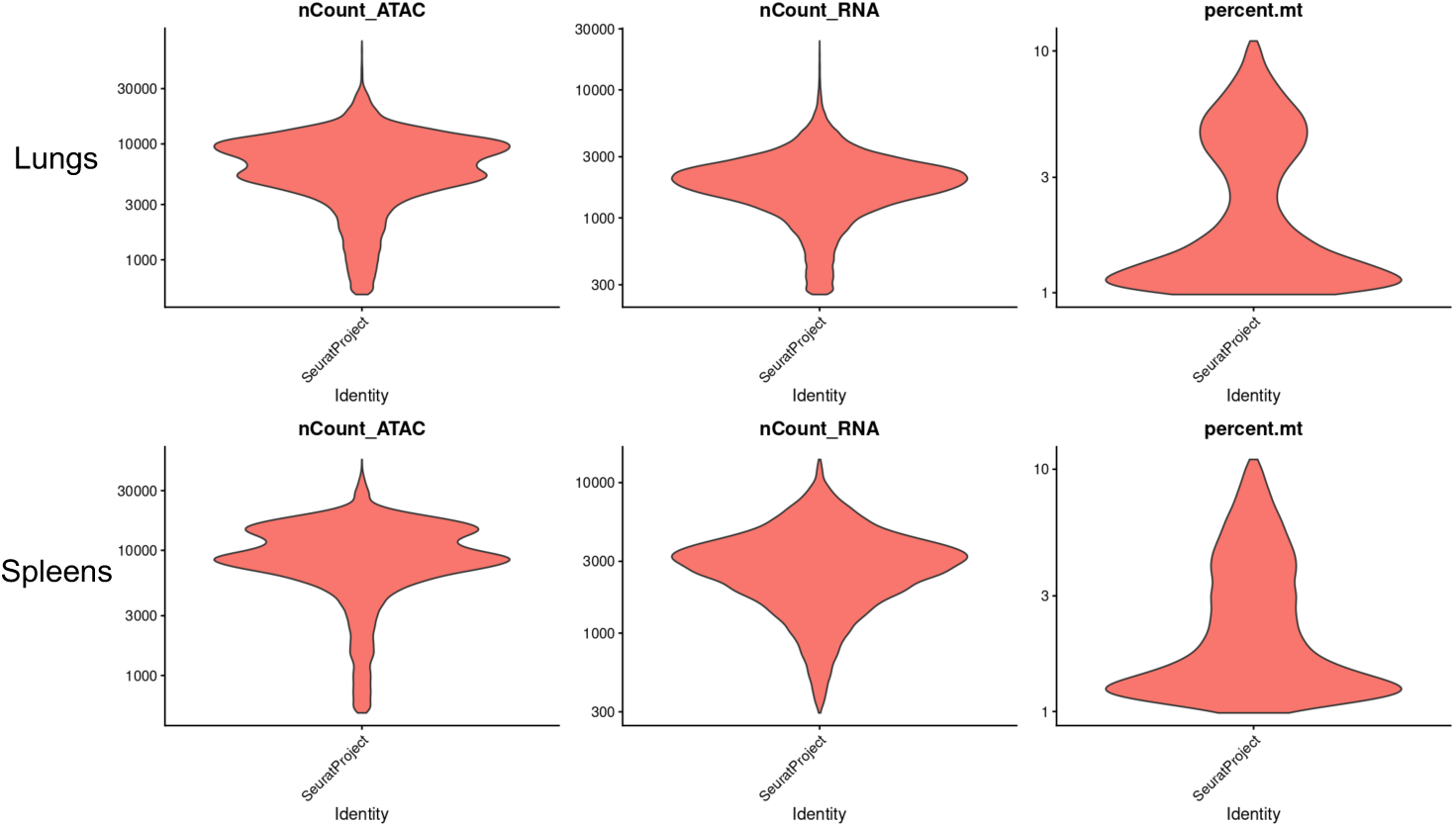
QC summary of single-cell multiomics libraries. Distribution of three QC quantities for lungs (top) and spleens (bottom) - read counts for ATAC libraries, read counts for RNA libraries, percent of mitochondria reads.

**Figure S2:**
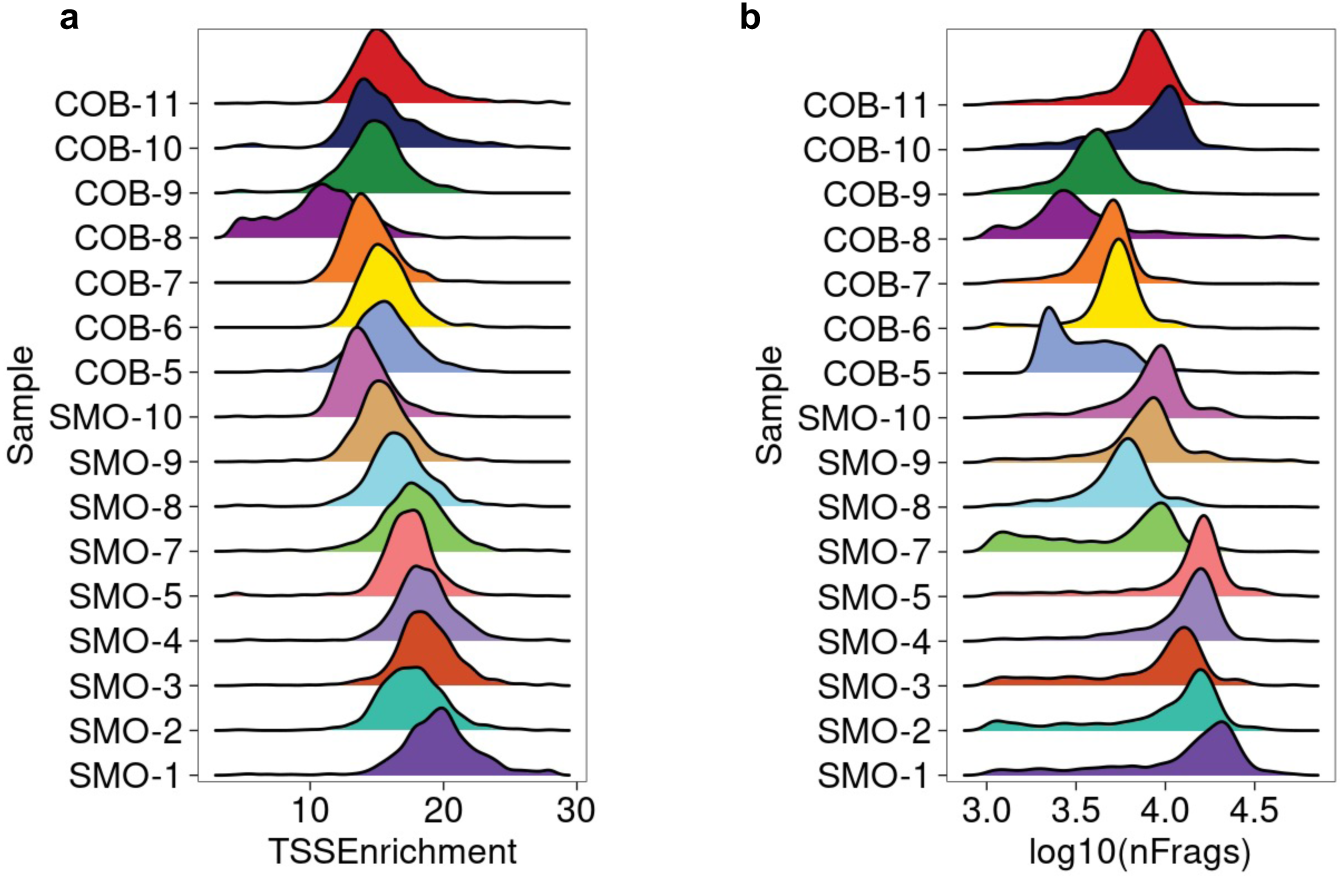
Sample QC for ATAC-seq libraries. The Y-axes for both figures denotes library IDs, which were divided into two large batches starting with “SMO” or “COB”. **a** Enrichment of ATAC-seq reads in transcript start sites. **b** Number of ATAC-seq fragments in the log scale

**Figure S3:**
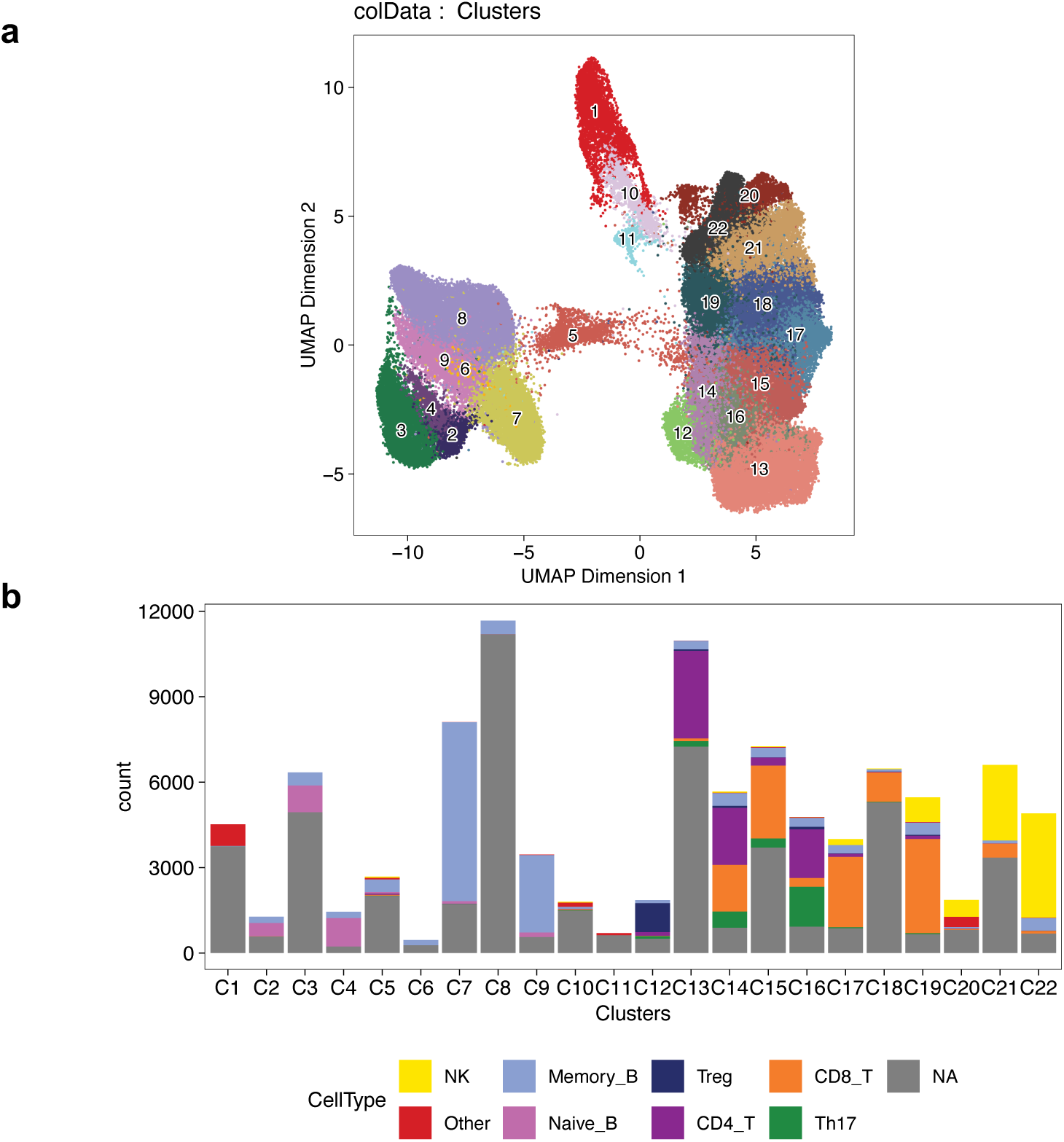
Clustering of pooled cells from both tissues using ATAC-seq data. **a**, UMAP visualization of cell clustering. **b**, The proportions of cells in the cell clusters that come from the mached RNA-seq libraries. The color scheme is consistent with Fig. 1b, except for cells that lack matched RNA-seq libraries (labeled as NA) in grey.

**Figure S4:**
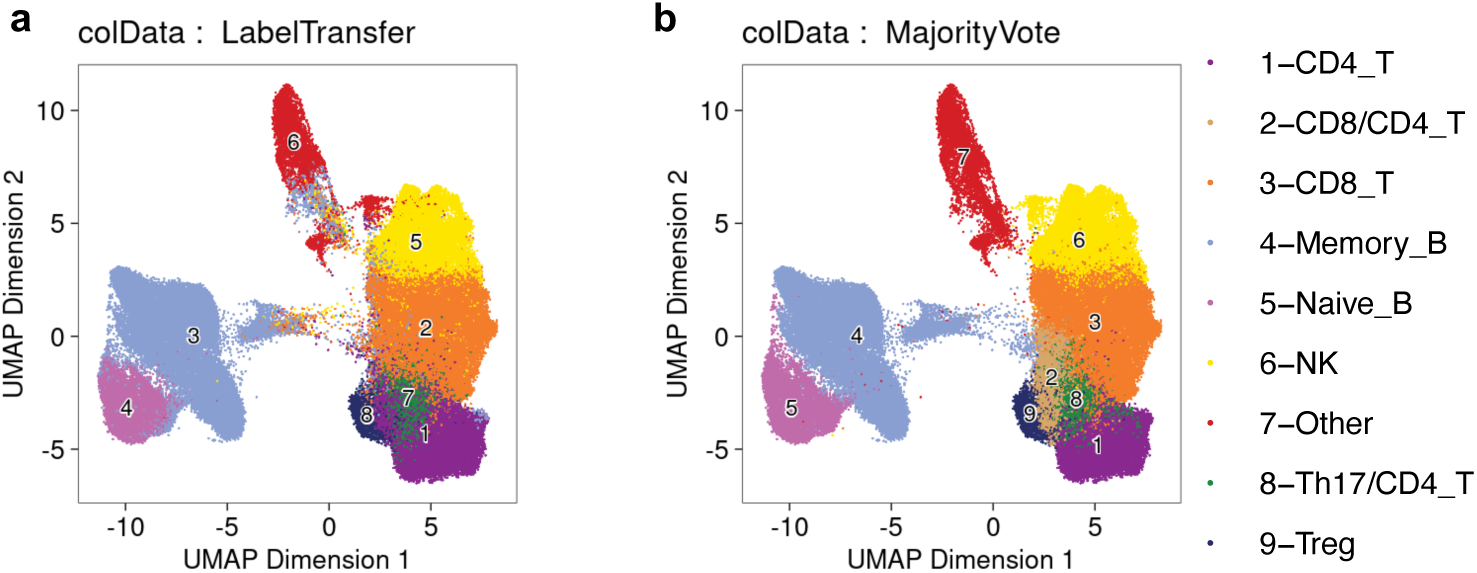
Comparison of two methods on assigning cell types for ATAC-seq libraries. **a**, label transferring from Seurat, **b**, majority-voting.

**Figure S5:**
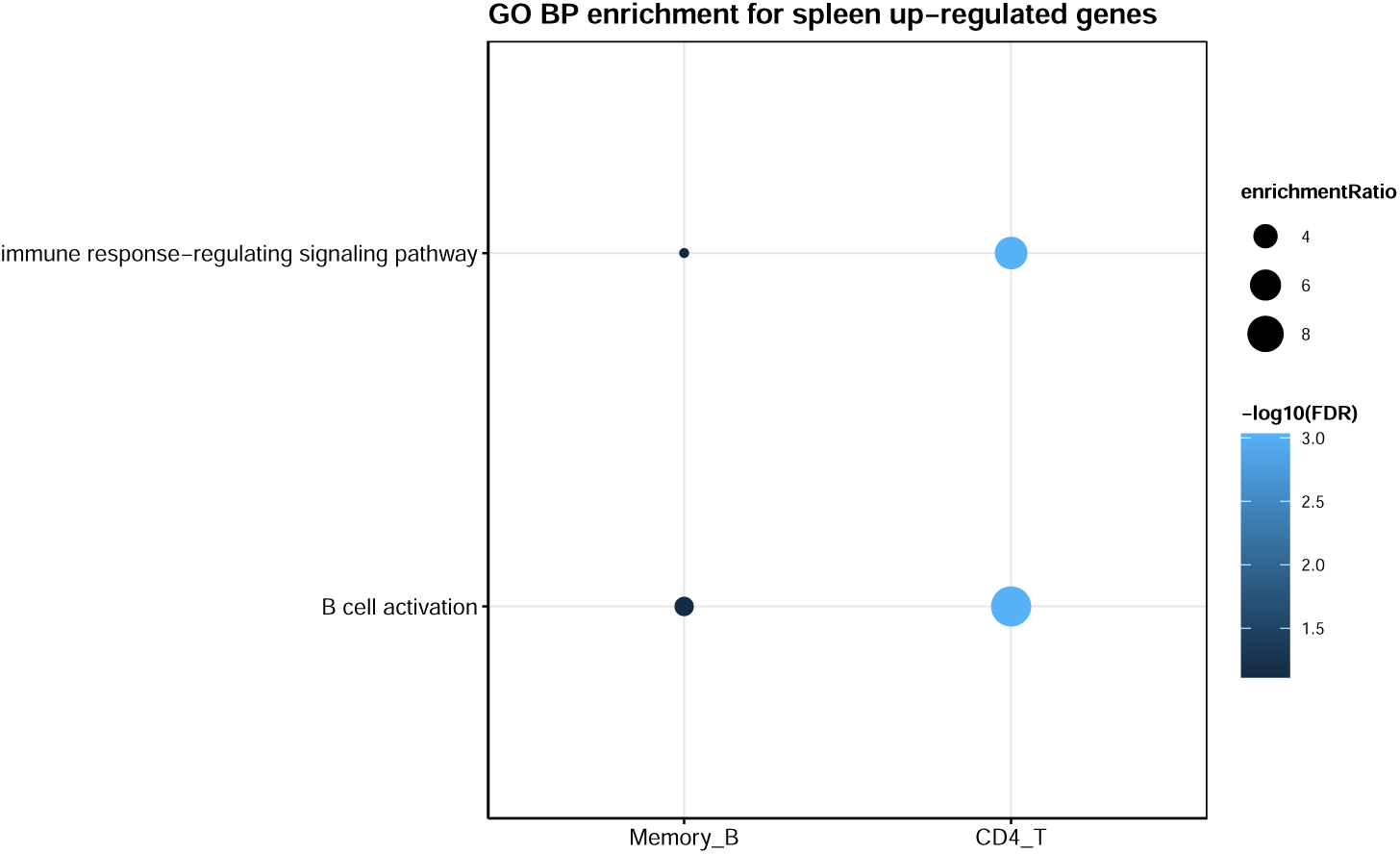
Top gene ontology terms enriched for spleen up-regulated genes in each cell type. Bubble size denotes fold of enrichment, while color denotes -log10(FDR).

**Figure S6:**
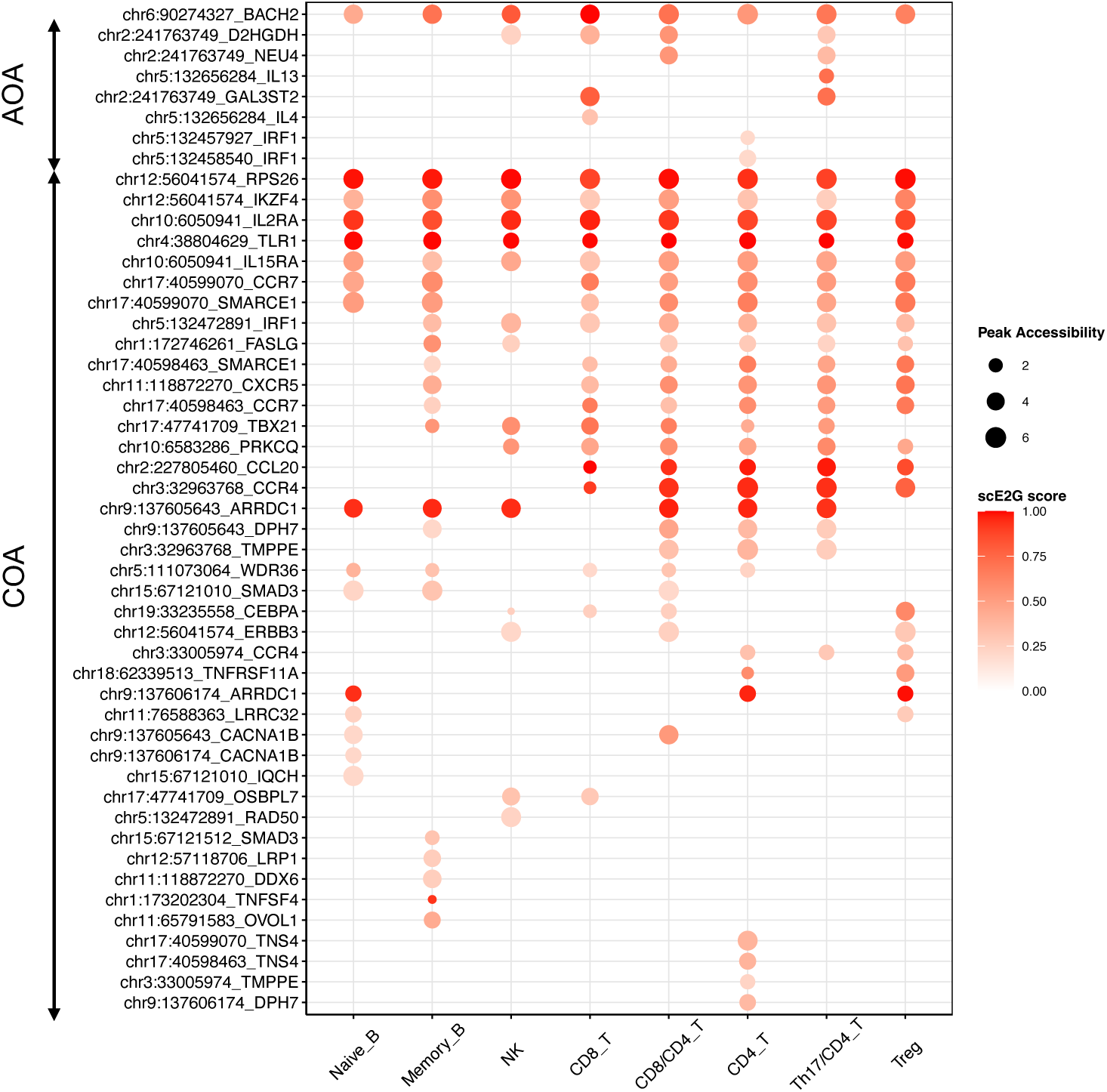
Bubble plot for the full list of CRE-gene pairs. The size of the bubble denotes absolute peak accessibility and the color indicates scE2G prediction scores. CREs for adulthood on-set asthma are on the top and those for childhood on-set asthma are at the bottom

**Figure S7:**
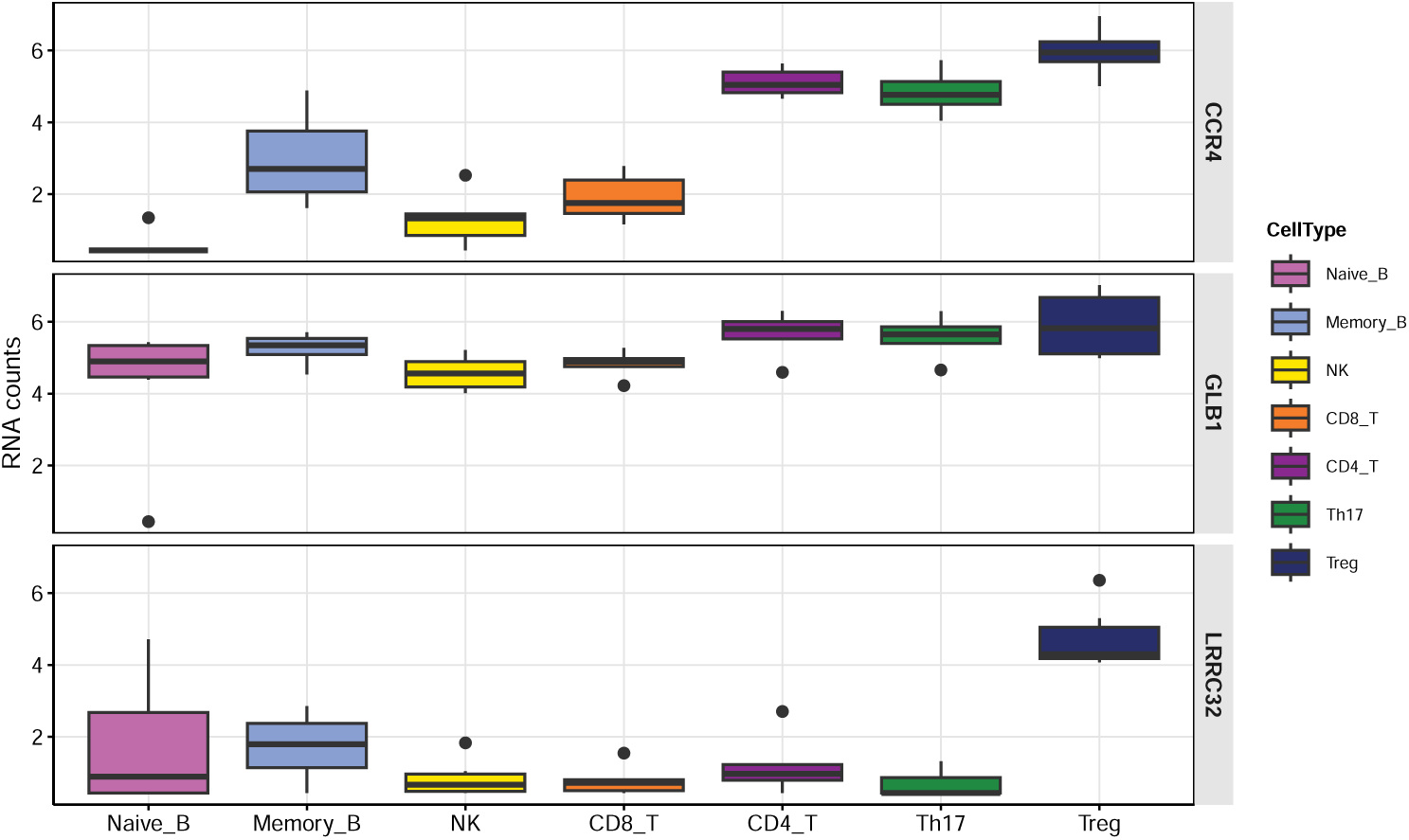
Pseudobulk RNA-seq read counts for *CCR4*, *GLB1*, and *LRRC32* across lung immune cell types. Comparing pseudobulk-level RNA-seq read counts for three genes shown in Figure 5 across lung immune cell types. The color of the boxplot represents different cell types.

**Figure S8:**
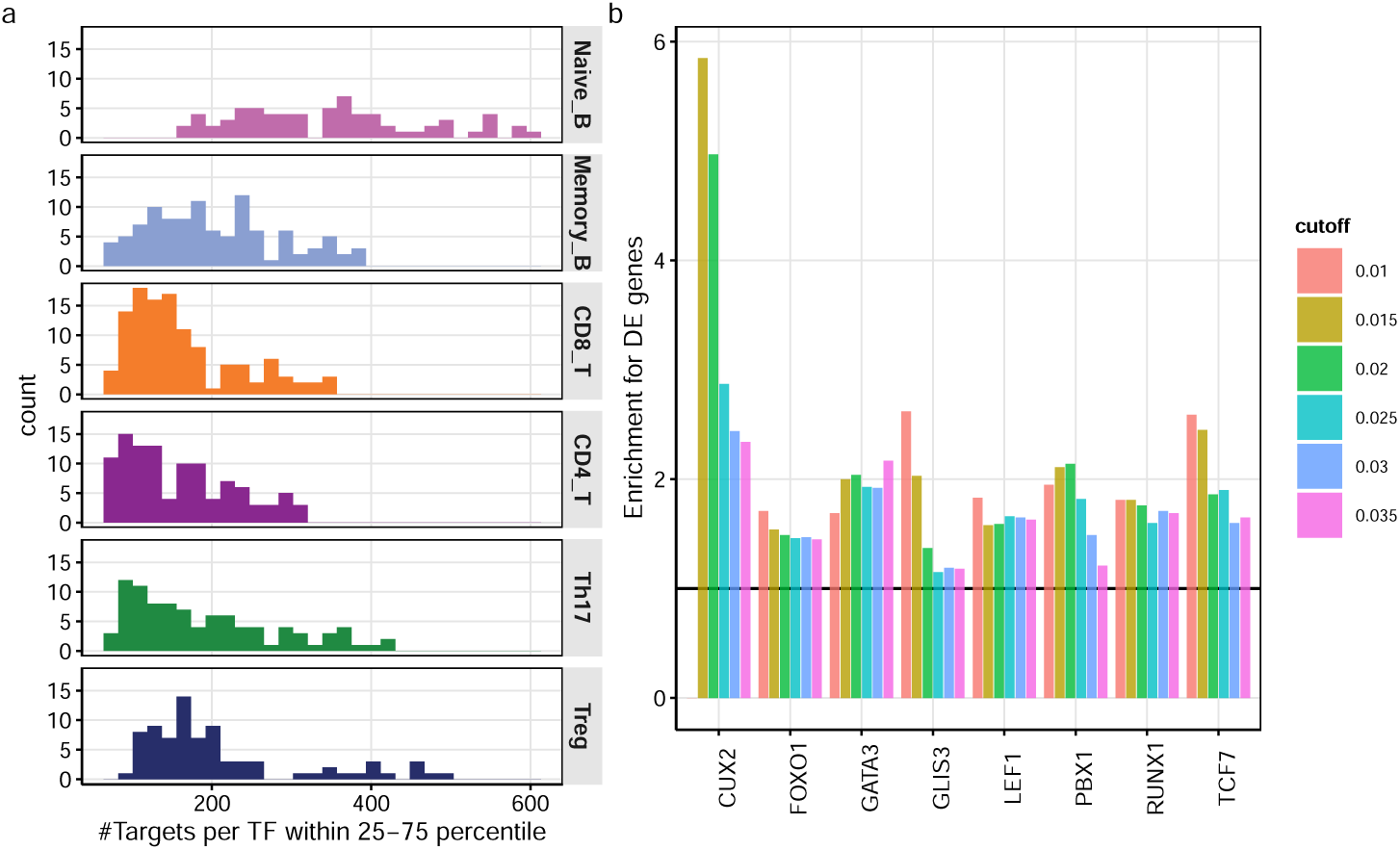
Characterization and validation of lung GRNs. **a**, Histogram for 25-75 percentile of the number of target genes per TF across cell types. **b**, A list of regulatory TFs of CD4 T cells whose target genes are significantly enriched for genes differentially expressed between CD4 T and memory B cells. The enrichment is assessed at different cutoffs of sparsity, a parameter used in defining the target genes of TFs.

**Figure S9:**
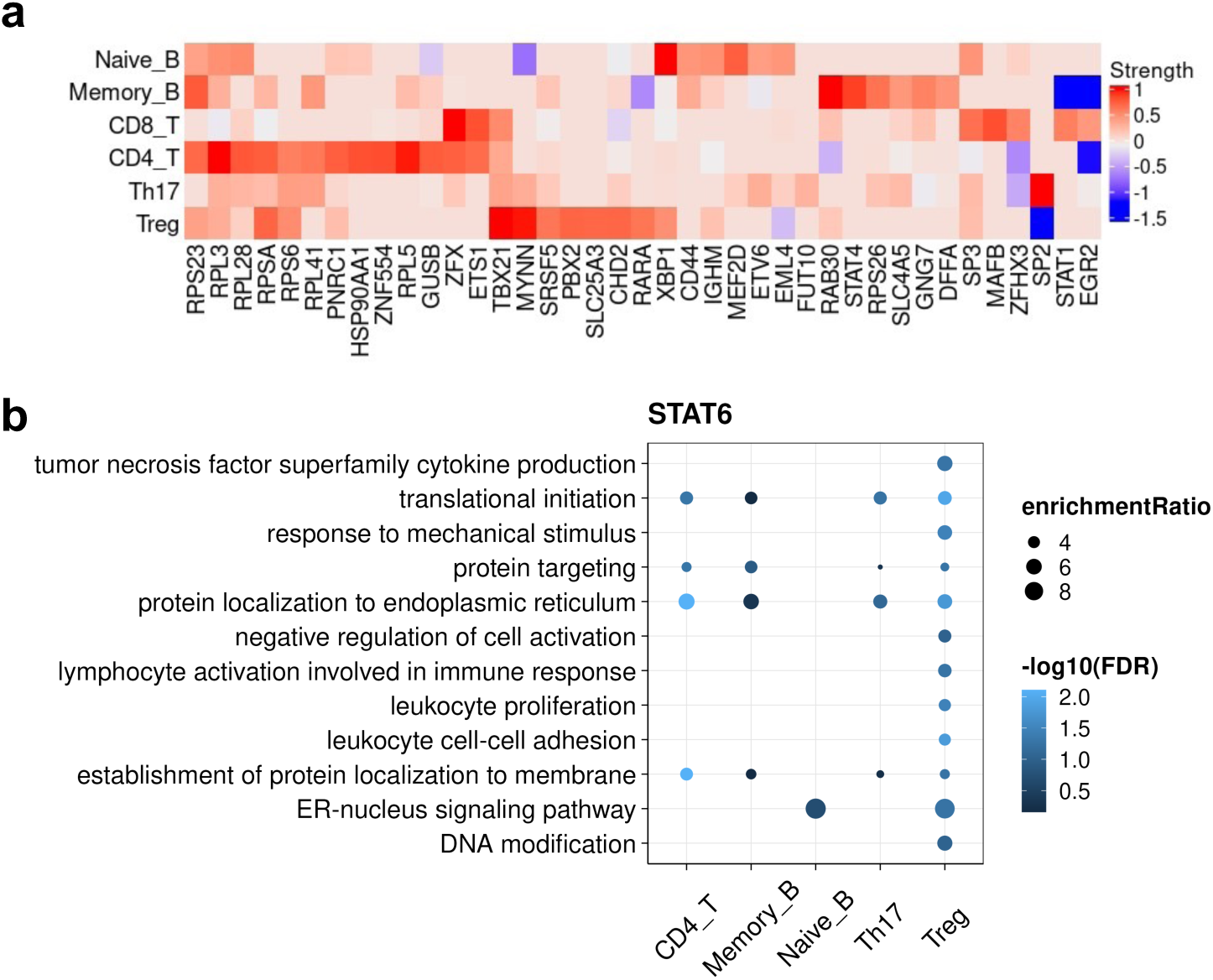
STAT6 target genes and their potential functions across cell types. **a**, Heatmap for *STAT6* regulation strength on target genes in different cell types. The color of regulation strength from red to blue represents positive to negative regulation. **b**, Top GO biological process terms enriched in STAT6 target genes in each cell type. Bubble size denotes fold of enrichment, while color denotes -log_10_(FDR).

**Figure S10:**
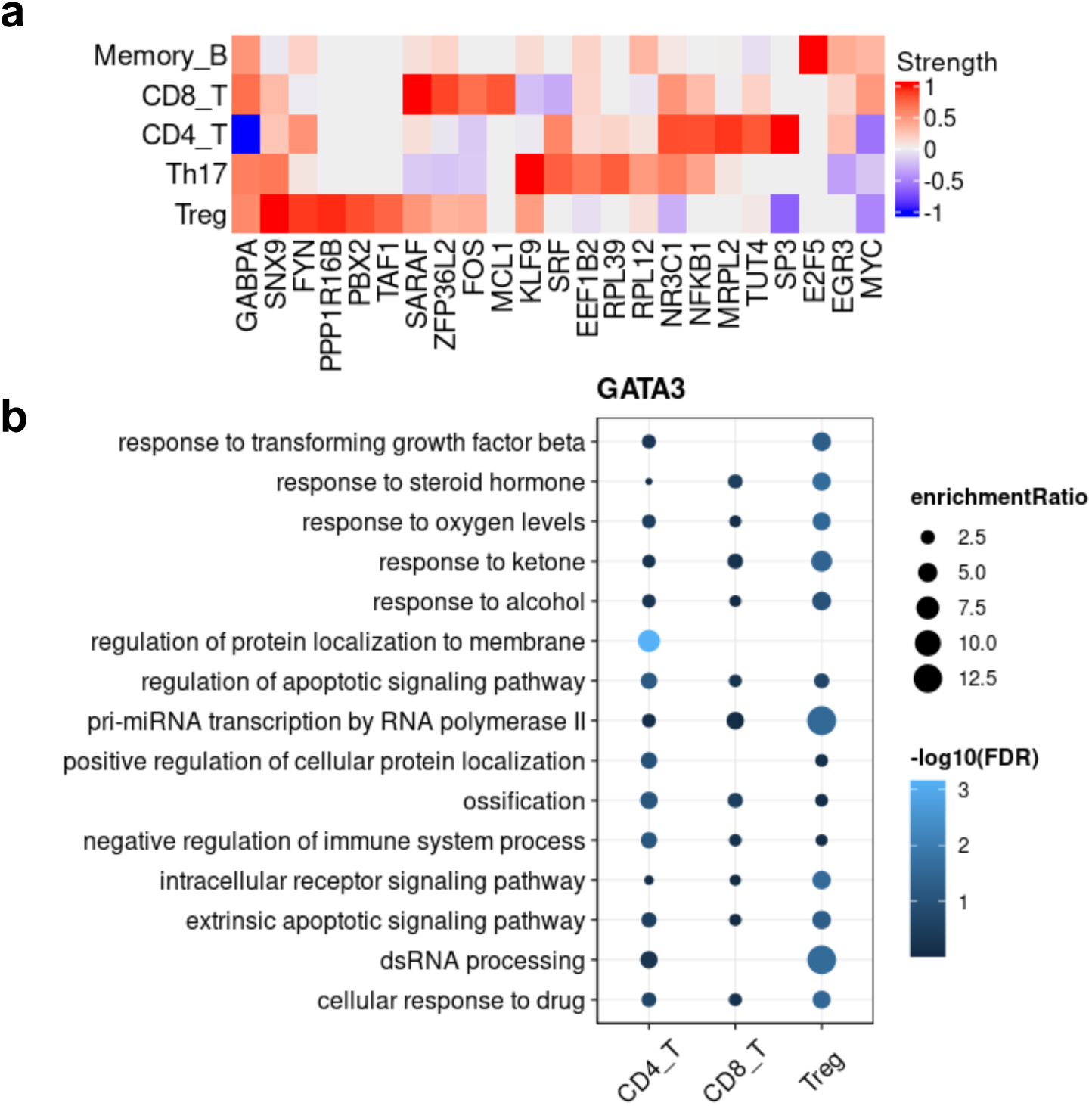
GATA3 target genes and their potential functions across cell types. **a**, Heatmap for *GATA3* the normalized regulation strength on target genes for different cell types. **b**, Top GO biological process terms enriched in GATA3 target genes in each cell type. Bubble size denotes fold of enrichment, while color denotes -log_10_(FDR).

**Figure S11:**
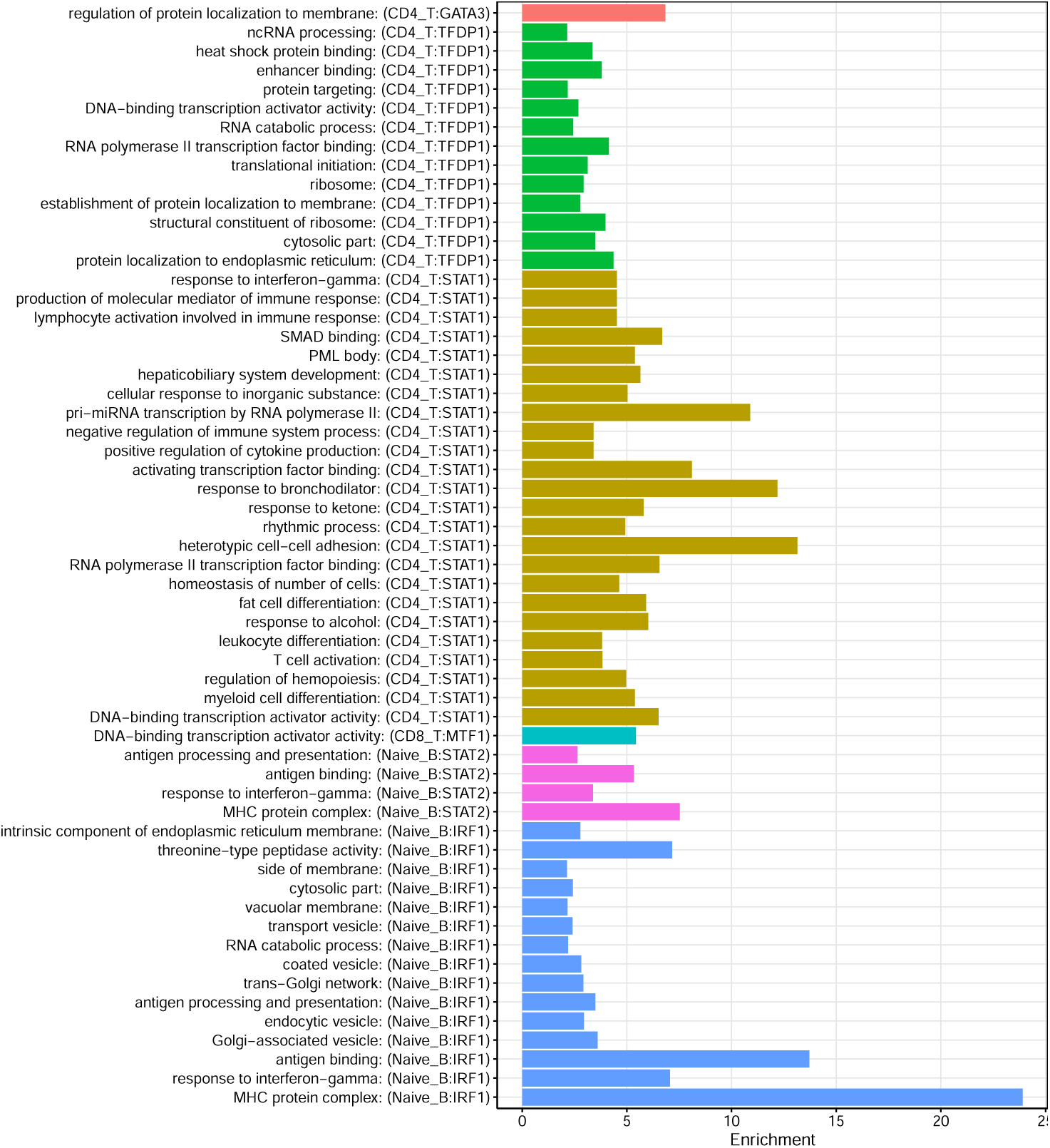
Full list of enriched GO biological processes terms for target genes of each candidate asthma TF in its respective cell type. Each label of Y-axis contains GO term, candidate TF and its respective cell type. The bar length represents the fold of enrichment for GO terms among the target genes. All the GO terms shown in the plot were above the cutoff of FDR at 0.05.

### Supplemental Tables

#### 5.1 Supplementary Tables

**Table S1: Summary of sample covariates for human lung and spleen tissues in this study**

**Table S2: Differentially expressed genes between tissue for each immune cell type .**

**Table S3: Enriched GO biological processes terms for lung up-regulated genes in individual immune cell types**

**Table S4: Enriched GO biological processes terms for spleen up-regulated genes in individual immune cell types**

**Table S5: Summary of differentially accessible peaks between tissue and between cell types**

**Table S6: Summary of scE2G predicted target genes using multiple annotations including PoPS scores**

**Table S7: Summary of predicted E2G links for the asthma-related CREs prioritized using our lung datasets**

